# Investigating the pathogenicity of missense spliceogenic variants using whole-genome sequencing, RNA-sequencing, and proteomics data from population and clinical research cohorts

**DOI:** 10.64898/2026.10.09.26365070

**Authors:** Mwenda R. Rintari, V. Kartik Chundru, Carolina J. Oquendo, Joseph Leslie, Alistair T. Pagnamenta, Diana Baralle, Caroline F. Wright

## Abstract

Missense variant pathogenicity is usually interpreted according to the predicted effect on the protein via polypeptide alterations. But missense variants can also cause disease by disrupting RNA splicing, here defined as missense spliceogenic variants (MSVs). Although splice prediction is increasingly incorporated into variant classification pipelines, it is not a standardised process, so the scale and clinical relevance of MSVs remains poorly defined. We integrated whole-genome sequencing and circulating proteomic data from ∼50,000 UK Biobank participants to characterise MSVs using ∼8.3 million missense variants. We found that 2.7% of missense variants are predicted to alter splicing (SpliceAIΔ ≥ 0.2), comparable with synonymous variants (2.8%). Notably, 57% had benign/neutral AlphaMissense and REVEL scores (<0.34 and <0.5 respectively), rising to 85.2% in intrinsically disordered regions, indicating that many MSVs would not be prioritised by protein-centric predictors.

MSVs were associated with reduced circulating levels of the cognate protein, approaching the effects of predicted loss-of-function variants. Analysis of raw SpliceAI scores revealed that events where novel cryptic sites outscored canonical sites resulted in the largest protein depletion, particularly when a premature frameshift was introduced. Validation of 14 predicted MSVs of uncertain significance in a clinical cohort provided strong pathogenic/likely pathogenic evidence, and screening unsolved cases from the 100,000 Genomes Project identified two novel candidate diagnostic MSVs. These findings establish the scale and functional consequences of MSVs, and demonstrate that RNA-centric interpretation can reveal clinically-relevant loss-of-function mechanisms missed by protein-centric missense variant interpretation.

## Introduction

Missense variants are a major contributor to human genetic disorders, accounting for approximately 38% of Mendelian conditions^1,2^. Traditionally, these variants are evaluated based on the effect of the predicted amino acid change and any functional or structural changes to the protein. However, as splicing machinery is dependent on the nucleotide sequence context at the pre-mRNA level, variants that are predicted to cause a missense change in the protein can also alter RNA splicing prior to translation, potentially resulting in complete loss-of-function through nonsense mediated decay (NMD)^3^; we term these missense spliceogenic variants (MSVs).

Literature estimates regarding the prevalence of MSVs mostly predate large-scale population genetic data and vary dramatically, ranging from 1% to 25%^4–8^. This wide discrepancy stems directly from differences in computational scope, regulatory motif definitions, and study cohort selection. Early database-wide screens focusing strictly on the loss of canonical splice-junction signals estimated that only 1–2% of disease-associated missense variants alter mRNA splicing^4,5^. Conversely, subsequent structural and biochemical analyses modelling the disruption of exonic splicing enhancers (ESEs) and creation of exonic splicing silencers (ESSs) suggested that 22–25% of inherited disease-causing variants alter functional exonic splicing signals^6,7^. Similarly, targeted clinical evaluations of gene-specific datasets, such as mismatch repair gene variants, have identified splice defects in up to 21% of evaluated missense variants^8^.

While variant analysis workflows routinely evaluate predicted synonymous variants for potential splicing disruptions^9^, missense variants rarely undergo equivalent systematic transcript-level evaluation due to their presumed coding consequences. Consequently, the field lacks a baseline estimation of this phenomenon at a population scale. By establishing this baseline, we can begin to evaluate exonic variants holistically, capturing pathogenic transcript-level aberrations as well as protein-level alterations, providing mechanistic insights that can aid variant interpretation.

Missense variants are often challenging to interpret, and currently represent ∼69% of all variants of uncertain significance (VUSs) in ClinVar^10^ (**Methods**). Clinical variant interpretation guidelines – such as those from the American College of Medical Genetics and Genomics (ACMG)^9^ and Association for Clinical Genomic Science (ACGS)^11^ – advise using tools such as AlphaMissense^12^ and REVEL^13^ to help inform on the prioritisation of missense variants, contributing to their *in-silico* evidence for pathogenicity. Tools such as these are generally protein-centric and are predominantly trained on residue-level evolutionary conservation and protein structure data. As a result, they are not optimised for predicting transcript-level effects, resulting in a potential blind spot in variant interpretation. Utilising transcript-centric tools such as SpliceAI^14^, Pangolin^15^, or MaxEntScan^16–18^ alongside protein-centric tools during missense variant interpretation may aid in resolving VUSs and identifying novel pathogenic missense variants.

Here, we present a systematic computational framework that integrates splice-prediction models with protein-centric variant effect predictions to capture MSVs that may result in loss-of-function through NMD. We leverage population-scale whole-genome sequencing (WGS) and plasma proteomic data from ∼500,000 and ∼55,000 UK Biobank (UKB) participants, respectively, to establish the first empirical genome-wide baseline of predicted MSVs. To demonstrate biological validity, we apply this to patient-derived whole-blood RNA-sequencing (RNA-seq) data from the 100,000 Genomes Project (100kGP), confirming the framework accurately identifies cryptic splice-site activations in missense variants previously classified as VUSs. Alongside this, we conduct a retrospective analysis of the Deciphering Developmental Disorders (DDD) study and the 100kGP to highlight several examples of pathogenic MSVs likely to act via splice disruption. Additionally, we functionally validate aberrant splicing as the pathomechanism for 14 clinically ascertained VUS from the NHS Genomic Medicine Service. Finally, we replicate our prioritisation workflow using unsolved 100kGP cases, successfully identifying the causative variant in a long-standing unsolved case and uncovering a strong candidate variant in two additional unsolved cases.

## Material and Methods

### Study Cohorts

#### UK Biobank Whole-Genome Sequencing and Proteomic Data

The UK Biobank (UKB) is a large-scale, prospective, population cohort containing genetic, lifestyle and clinical health data from ∼500,000 UK participants between 40-69 years of age at recruitment (2006-2010)^19^. The UKB Consortium performed sequencing using Illumina NovaSeq 6000 sequencing machines to generate 150-bp paired end reads, and called variants using DRAGEN v3.7.8^20^. We analysed WGS data from 490,640 UK Biobank (UKB) participants (GRCh38; average coverage 32.5x). To examine the functional consequences of these variants, we used circulating plasma proteomic data generated via Olink Proximity Extension Assay (PEA) technology by the UKB Pharma Proteomics Project (UKB-PPP) on 2,923 proteins from 54,219 UKB participants^21^.

#### The 100,000 Genomes Project Cohort and Transcriptomic Data

The 100,000 Genomes Project (100kGP), led by Genomics England, is a large-scale national genomic medicine initiative that includes integration of clinical, whole-genome sequencing data^22,23^. Whole-blood RNA sequencing data from >7000 rare-disease patients and/or family members is available, and methods used to generate these transcriptomes was described previously^24^.

#### The Deciphering Developmental Disorders Study

The Deciphering Developmental Disorders (DDD) study is a UK-wide translational research study of >13,500 families designed to identify the genetic causes of undiagnosed, severe developmental disorders in children using exome sequencing, microarrays and clinical phenotyping^25^. Clinically-assessed pathogenic/likely pathogenic variants are available via DECIPHER^26^.

#### The NHS Genomic Medicine Service

The NHS Genomic Medicine Service (NHS GMS) is the national clinical diagnostic service in England for high-throughput genomic testing of patients with rare diseases. Clinical testing and patient eligibility are governed by the National Genomic Test Directory, with diagnostic validations performed in UKAS-accredited clinical laboratories. All individuals included in this study were identified through NHS clinical referral pathways following the detection of variants of unknown significance considered by the referring clinician to be plausible explanations for the patient’s phenotype. No additional pre-screening criteria were applied.

The majority of participants were enrolled in the University of Southampton Splicing and Disease study (IRAS 49685), enabling access to detailed phenotypic and clinical information. A small number of additional cases were referred directly through NHS diagnostic pathways for functional assessment of candidate spliceogenic variants and no phenotypic data is available.

### Variant Prioritisation and *In-Silico* Prediction

#### Identification and Enhanced Annotation of Missense Spliceogenic Variants

We define missense spliceogenic variants (MSVs) as variants with a predicted missense consequence that are also predicted to disrupt canonical pre-mRNA splicing in the same transcript to produce abnormally spliced mRNA. To identify predicted MSVs, we annotated all single nucleotide variants (SNVs, n=1,217,604,791) in the UKB WGS data using the Ensembl Variant Effect Predictor (VEP, v.110)^27^, in conjunction with the Loss-Of-Function Transcript Effect Estimator (LOFTEE) plugin^28^. We restricted downstream analyses to variants predicted to have a missense or synonymous consequence in MANE Select (v1) transcripts^29^ (missense n=8,323,258; synonymous n=3,753,457). We excluded 909 variants across eight genes (*ACYP2*, *IL36B*, *TSTD3*, *UFSP1*, *RGS20*, *SMIM2*, *PRY*, and *PRY2*) because updates to their MANE Select transcript assignments prevented SpliceAI annotation. We further annotated missense variants with AlphaMissense^12^ and REVEL^13^ metapredictors, and used these scores to select variants that were predicted to be tolerated (AlphaMissense <0.34; REVEL <0.5) or damaging at the protein level **(Supplementary Table S1)**. Allele frequencies for UKB variants were obtained from gnomAD v4.1.

To predict the splicing impact of these variants, we utilised SpliceAI, a deep residual neural network that evaluates sequence context to calculate the probability of any given base in a given window acting as a splice acceptor or donor site^14^. To classify the predicted impact, we implemented the three standard delta score thresholds between wild-type and variant, established by Jaganathan et al: a sensitive threshold (Δ ≥ 0.2), a moderate-confidence recommended threshold (Δ ≥ 0.5), and a high-precision threshold (Δ ≥ 0.8) **(Supplementary Table S1)**. All missense and synonymous variants were annotated using SpliceAI against MANE Select (v1.4) with an extended flanking window of 500 basepairs (bp). To this end, we updated the SpliceAI source code for compatibility with current versions of tensorflow, and for computational efficiency we reduced the floating point precision of the models from fp32 to fp16 (https://github.com/chundruv/SpliceAImod). In addition to the compatibility updates and optimisations, we also include extra inference on the splice sites including the predicted splice event (frameshift vs inframe insertion or deletion), comparison of the raw probabilities of the Δposition site to the canonical splice site, and the identification of multiple predicted splice events.

Missense variants observed with high SpliceAI scores (Δ ≥ 0.2) and low protein-centric scores (AlphaMissense <0.34 and REVEL <0.5) are termed Low Missense-scoring Splice Variants (LMSVs).

#### Splice-Junction Mapping

To establish the positional context of our variants, we developed a custom, strand-aware splice-junction reference using GENCODE^31^ (v48) basic annotations restricted to MANE Select (v1.4) multi-exon transcripts (github.com/MRintari/Missense-Spliceogenic-Variant-Analysis). Using this reference annotation file, we calculated the physical distance in base pairs between each UKB missense variant and its closest canonical splice junction, and classified the proximal junction as either a splice donor or a splice acceptor.

#### Extending SpliceAI Masking Quality Control to Distal Splice-junctions

To address a known limitation in SpliceAI’s masking over-extended windows^30^, where gain events at the proximal annotated splice site are scored at zero while occasionally leaving more distal sites in transcripts unmasked, we developed an extended masking quality control pipeline using our splice-junction reference file. We used this to systematically cross-reference every predicted splice-gain event from our candidate variants (LMSVs) against these annotated splice junctions to identify and filter out any unmasked distal artefacts.

#### Quantifying The Proteomic Consequences of Missense Spliceogenic Variants

To evaluate the downstream functional consequences of predicted MSVs on protein abundance, we analysed the UKB-PPP circulating plasma proteomic data^21^. We stratified variants in genes mapping to 963 plasma proteins into four primary groups across seven functional categories **(Supplementary Table S1)**:

- **Benchmark Loss-of-Function Groups**: We established a positive control benchmark for severe protein depletion comprising predicted Loss-of-Function (p.LoF; n = 1,698) variants and experimentally validated splice-altering variants (n = 143) from SpliceVarDB^33^. We defined p.LoF single nucleotide variants (SNVs) strictly using high-confidence LOFTEE predictions, a high-impact classification from Ensembl VEP, a CADD^34,35^ score ≥ 28.1, and a high-impact classification from Ensembl VEP. High-impact classifications cover transcript ablation, splice acceptor, splice donor, stop gain, frameshift, stop loss, start loss, transcript amplification, feature elongation, and feature truncation variants. We restricted the SpliceVarDB benchmark to variants with splice-altering evidence across Multiplexed Functional Assays of Splicing using Sort-Seq (MFASS), Massively Parallel Splicing Assays (MaPSy), RNA-sequencing (RNA-seq), or minigene reporter assays.
- **Predicted Splice-Driven Groups**: We selected predicted MSVs (p.Splice Missense; n = 392) using a high-precision SpliceAI threshold (Δ ≥ 0.8) paired with low protein-centric scores (REVEL < 0.5; AlphaMissense < 0.34); we term these high-confidence LMSVs). Predicted synonymous spliceogenic variants (p.Splice Synonymous; n = 449) were defined using an identical SpliceAIΔ ≥ 0.8 threshold.
- **Predicted Missense-Driven Group**: We curated a p.Missense Damaging category (n = 1,407) by selecting variants with negligible predicted spliceogenic impact (SpliceAIΔ ≤ 0.01) paired with high-confidence pathogenic protein-centric scores (REVEL ≥ 0.932; AlphaMissense ≥ 0.564).
- **Control Groups**: We defined negative control benchmarks by requiring negligible predicted splicing disruption (SpliceAIΔ ≤ 0.01) and a CADD score threshold of ≤ 0.15. These consisted of Synonymous Controls (n = 8,722) and Missense Controls (n = 992), which required predicted tolerated protein-centric scores (REVEL < 0.5; AlphaMissense < 0.34).

We adjusted our association analyses for age, age^2^, sex, WGS batch, Olink plate, fasting time, UKB recruitment centre (as a proxy for geography), the first 40 genetic principal components, and the time since blood draw. We rank-inverse normalised circulating protein level measurements at runtime.^36^

We adjusted p-values for multiple testing using the Benjamini-Hochberg false discovery rate (FDR) method with a significance threshold of q < 0.05 to identify significant variant-protein association effects^37^. We evaluated pairwise differences in effect size (◻) distributions between the variant categories using non-parametric, two-sided permutation tests executed with 50,000 random permutations. We subsequently corrected the resulting permutation p-values using the Benjamini-Hochberg procedure.

Precomputed FoldX thermodynamic stability scores (ΔΔG) for the p.Splice Missense and p.Missense Damaging groups were downloaded from the EMBL-EBI ProtVar repository (accessed February 2026)^38,39^. We mapped residue-level FoldX predictions to variants using UniProt protein coordinates; 174 of the 1,797 variants failed to map and were excluded from downstream structural analysis. Finally, we applied a thermodynamic threshold of ΔΔG < 2 kcal/mol to obtain a high-confidence subset of missense variants predicted to maintain wild-type structural conformation **(Supplementary Table S1)**.

#### Prediction of Intrinsically Disordered Regions

Data downloaded from the EMBL-EBI ProtVar repository included AlphaFold predicted local distance difference test (pLDDT) scores, which we mapped to unique residues corresponding to missense variants across UKB MANE Select transcripts (N = 3,154,276 residues). Residues were categorised into predicted intrinsically disordered regions (IDRs; pLDDT < 50) or predicted structured regions (pLDDT ≥ 50) based on standard confidence pLDDT thresholds^40^. To control for potential terminal positioning bias (such as N- or C-terminal enrichment of disorder or reduced splice disruption), sensitivity analyses were conducted by restricting the residue dataset to the central 70% of each protein. Proportional differences in splice disruption baseline frequencies and LMSV frequency were evaluated using two-sided Fisher’s exact tests to obtain Odds Ratios (OR) and corresponding p-values.

### Splicing Validation using RNA sequencing data

#### Validation of Spliceogenic Variant Effects on RNA Splicing using Clinical Cohorts

To validate the biological accuracy and clinical relevance of our computational prioritisations at the transcript level, we reproduced our prioritisation framework with clinical transcriptomic datasets. First, we downloaded all ClinVar variant data available (accessed August 2025) and annotated all variants using SpliceAI with a 500-bp sequence context as previously detailed. We used this data to determine the percentage of VUSs that are missense variants. Additionally, we calculated the distribution of MSVs across clinical significance tiers using thresholds of Δ ≥ 0.2, ≥ 0.5, and ≥ 0.8.

We restricted downstream prioritisation to variants of uncertain significance (VUSs) within genes holding high-confidence (green status) associations with developmental disorders in PanelApp, yielding 537,127 candidate VUSs. We identified LMSVs by requiring a SpliceAIΔ ≥ 0.2 alongside concordantly benign or neutral protein-centric predictions (REVEL < 0.5 and AlphaMissense < 0.34), resulting in 6,709 candidate VUS-LMSVs **(Supplementary Table S1)**.

We intersected these VUS-LMSVs with the 100kGP tiered variant database and identified carrier participants with available whole-blood RNA-sequencing (RNA-seq) data (accessed via LabKey February 2026). We retained carriers where the target variant locus and adjacent canonical splice junctions achieved a total read depth across carriers of ≥ 2 and the cryptic splice site demonstrated a change in Percent Spliced In (ΔPSI) of ≥ 15% relative to their control group (ΔPSI = PSI_carrier_ - PSI_controls_). We screened for candidate splice disruptions using the Integrative Genomics Viewer (IGV). As all assessed carriers were heterozygous, we performed allele-specific read analysis by grouping reads according to the nucleotide present at the variant position. This allowed us to directly link aberrant splicing events to the variant-read and distinguish real splice disruptions from background transcript variation. To minimise technical artefacts and batch effects stemming from sequencing and alignment, control individuals were selected from the same Illumina delivery batch as the respective carrier probands. We statistically evaluated differences in cryptic splice-site usage between carrier individuals and delivery batch-matched controls using a two-tailed Fisher’s exact test (p < 0.05).

VUS-LMSVs disrupting splicing via intron retention were assessed using custom Python scripts to quantify allele-specific exon-intron boundary coverage from RNA-seq BAM files (github.com/MRintari/Missense-Spliceogenic-Variant-Analysis). High-quality reads (mapping quality score ≥ 20, excluding duplicate, secondary, and supplementary reads) spanning the targeted splice-junction were evaluated, with reads extending ≥ 2bp across both sides of the boundary being classified as demonstrating intron retention. Statistical significance of intron retention was assessed through a complementary intra- and inter-sample analysis. Within carriers, we performed an intra-sample allele-specific analysis using a two-tailed Fisher’s exact test to compare the ratio of canonically spliced reads to intron-retaining reads between alternate and reference alleles. For inter-sample comparisons against control cohorts, we controlled for technical variability across sequencing and delivery runs using a Cochran-Mantel-Haenszel (CMH) test stratified by pipeline processing batch. Additionally, we modelled intron retention using a Linear Mixed-Effects Model with experimental group (carriers vs. controls) and delivery batch included as fixed effects, and sample ID as a random intercept.

#### Transcriptomic Validation of Clinical Referrals (NHS Genomic Medicine Service)

We identified probands with RNA data available that harboured variants with high SpliceAI scores (n=14). RNA-sequencing and RT-PCR analyses were performed as previously described^41^. Splicing abnormalities were assessed through visual inspection of aligned reads using Integrative Genome Viewer (IGV) by at least two independent scientists.

Transcriptomic data was reviewed to characterise the nature and consequences of the splicing defects, including exon skipping, cryptic splice-site usage, and intron retention where present. Formal variant classification was not undertaken.

### Clinical Application and Diagnostic Utility

#### Retrospective Diagnostic Assessment of Solved Cases in Clinical Cohorts

##### 100kGP

We obtained genomic and clinical records from 9,046 solved probands within the 100kGP cohort. We filtered these solved cases to isolate tiered, diagnostic variants that matched our LMSV criteria, identifying 15 candidate cases. For participants whose reported causal variant in the clinical exit questionnaire matched one of the 15 identified LMSVs, we manually reviewed the accompanying comments. We screened these candidate variants for presence among unaffected 100kGP cohort participants, and obtained Pangolin scores alongside SpliceAI scores. For all variants satisfying these criteria, we cross-referenced corresponding records in the ClinVar database which also provide functional validation data or previously published evidence of splice disruption.

##### DDD Study

We selected clinically-classified pathogenic/likely pathogenic missense variants identified as diagnostic in the DDD study^25^ then filtered variants to only include LMSVs (predicted missense in MANE Select transcripts, SpliceAI ≥0.2, REVEL <0.5, AlphaMissense <0.34).

#### Application of Missense Spliceogenic Variant Prioritisation Workflow to Unsolved Cases in the 100kGP

We applied our computational prioritisation workflow to systematically screen unsolved cases within the 100kGP cohort, filtering for individuals who possessed a homozygous alternative genotype for any tiered LMSVs. We cross-referenced associated candidate genes against PanelApp, restricting our search to genes with high-confidence (green status) disease associations. For the remaining variants, we manually reviewed clinical records to confirm that the disease-association aligned with the proband’s recorded phenotype.

We further filtered candidate variants using the following criteria: a gnomAD allele frequency of < 0.01, fewer than three homozygous individuals recorded in gnomAD, and zero unaffected homozygous individuals across both the 100kGP and UKB cohorts. Where available, we examined read-level RNA-sequencing data using IGV software.

## Results

### Predicted Splicing Disruption Is Comparable Between Missense and Synonymous Variants and Is Not Identified by Protein-Centric Pathogenicity Predictors

We evaluated 8,323,258 missense and 3,753,457 synonymous variants within MANE Select transcripts from the UK Biobank (UKB) cohort. We found that 2.71% (n = 225,430) of missense variants possessed a SpliceAIΔ ≥ 0.2, which is comparable to 2.75% (n = 103,153) observed for synonymous variants (Cohen’s h = -0.002; **Figure 1A**). We observed this parity across the recommended and high-precision thresholds. Specifically, variants with a SpliceAIΔ ≥ 0.5 accounted for 0.82% (n = 68,177) of the missense variants compared to 0.79% (n = 29,639) of synonymous variants, while the most stringent threshold (SpliceAIΔ ≥ 0.8) captured 0.31% (n = 25,435) of missense and 0.29% (n = 10,812) of synonymous variants. Stratifying predictions by event type confirmed that score distributions align directly with canonical splice site motif architecture (**Supplementary Figure S1)**. Donor events exhibited higher overall score distributions compared to acceptor events, reflecting the greater importance of the exonic motif for donor recognition (three exonic bases vs one for acceptors), with acceptor loss showing a steep, exponential decay due to the single relevant exonic base.

**Figure 1:**
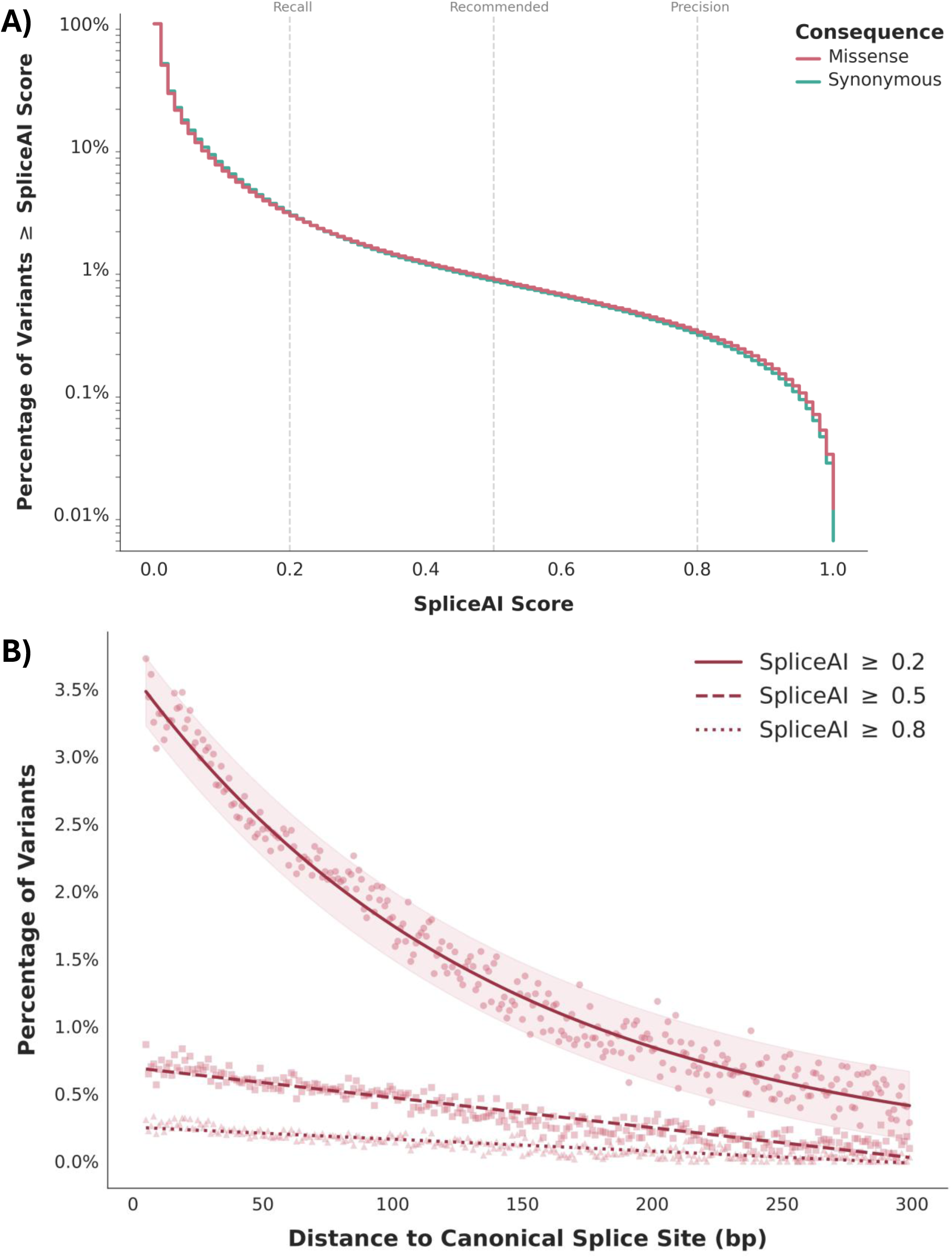
Prevalence and spatial distribution of predicted missense spliceogenic variants (MSVs). (A) Cumulative distribution of SpliceAIΔ scores for missense (red) and synonymous (teal) variants; vertical dashed lines denote standard thresholds for Recall (0.2), Recommended (0.5), and Precision (0.8) from SpliceAI. (B) Distribution of predicted MSVs relative to canonical splice sites; percentage of MANE Select UKB missense variants predicted to disrupt splicing, stratified by SpliceAI Δ thresholds: ≥ 0.2 (solid line, circles), ≥ 0.5 (dashed lines, squares), ≥ 0.8 (dotted line, triangles); distances reflect variants 5-300 base pairs away from a canonical splice junction.

We stratified the missense and synonymous variants across five gnomAD genomic allele frequencies (**Supplementary Table S2**), observing a steady decline in the prevalence of predicted spliceogenic variants as allele frequency increased across both predicted consequences. At the SpliceAIΔ ≥ 0.2 threshold, predicted splice-disrupting variants comprised 2.80% of missense variants not observed in gnomAD (v4.1.1) compared to 1.67% of common missense variants (AF > 1%). This depletion was more pronounced at high-confidence thresholds; variants with SpliceAIΔ ≥ 0.8 were over 3-fold less frequent among common missense variants (0.1%) compared to unobserved variants (0.33%). Parity between missense and synonymous variants was preserved across every frequency tier (**Supplementary Figure S2**), with a stratified Mantel-Haenszel test controlling for allele frequency confirming equivalence at each SpliceAI threshold (OR_mean_ = 1.01, p_mean_ = 8.03 × 10^−3^; **Supplementary Table S3**).

Among the 225,430 predicted MSVs with SpliceAIΔ ≥ 0.2, we found that 72% (n = 162,528) were classified as benign or neutral by at least one standard protein-centric tool, scoring either AlphaMisense < 0.34 or REVEL < 0.5 (**Supplementary Figure S3**). When restricting this subset to the 174,896 predicted MSVs where we successfully obtained scores from both predictors from Ensembl VEP, 82% score below the threshold for one of the two tools and 57.1% (n = 99,193) simultaneously scored below these thresholds on both tools.

### The Splicing Impact of Predicted Missense Variants Extends Deep into Exons

The predicted spliceogenic burden of missense variants decays in a distance-dependent manner relative to canonical splice junctions (**Figure 1B**). At canonical splice sites, we observed a stark contrast between donor and acceptor sites; 61% of missense variants at a donor position possessed a SpliceAIΔ ≥ 0.2 (26,193 / 42,946), whereas only 26.1% reached this threshold at an acceptor site (11,190 / 48,295) (Chi-square test, p < 2.2 × 10^-16^; **Supplementary Figure S4**). Although the highest density of predicted MSVs clusters within the canonical splice region, this elevated spliceogenic burden extends well into adjacent exonic regions. Specifically, 3.4% of missense variants located 5-19 bp away from a canonical splice site have a SpliceAIΔ ≥ 0.2 (39,991 / 1,193,412), significantly exceeding the genomic missense baseline observed across the rest of the genome (2.6%; 185,439 / 7,129,846) (Chi-square test, p < 2.2 × 10^-16^). This localised enrichment represents a 6.8-fold increase in predicted MSVs compared to variants located more than 300 bp away from a canonical splice junction (0.5%).

### Predicted Missense Spliceogenic Variants Result in Depleted Circulating Plasma Levels of Cognate Proteins in the UK Biobank

We assessed variant effects on the circulating level of cognate proteins in the UKB across 963 plasma proteins, which we stratified into four primary groups: two benchmark loss-of-function variants (p.LoF and experimentally validated splice variants^33^), predicted splice-driven variants (p.Splice Synonymous and high-confidence low missense-scoring splice variants [LMSVs]), a predicted missense-driven group (p.Missense Damaging), and two negative controls (predicted tolerated missense and synonymous variants; **Methods**). Here, LMSVs define missense variants with high splicing potential but low protein-centric pathogenicity scores, with high-confidence LMSVs prioritised using a strict SpliceAI threshold (Δ ≥ 0.8; REVEL < 0.5, AlphaMissense < 0.34). High-confidence LMSVs significantly reduced abundance of the cognate protein (◻_median_ = -0.61), approaching the severe depletion observed in the p.LoF benchmark (◻_median_ = -0.94; **Figure 2A**). This reduction in protein abundance was highly significant compared to the tolerated missense control (◻_median_ = -0.06, q = 1.1 × 10^-4^) and synonymous control (◻_median_ = 0.001, q = 1.1 × 10^-4^), both of which exerted a negligible effect on circulating cognate protein levels (**Figure 2B**). Furthermore, we detected no significant difference in protein abundance between HC-LMSVs and either the known validated splice variant benchmark (◻_median_ = - 0.34, q = 0.11) or p.Splice Synonymous variants (◻_median_ = -0.37, q = 0.11). Notably, we see no significant difference in abundance of the cognate protein between the HC-LMSVs and the p.Missense Damaging group (β_median_ = -0.71, q = 0.3956), likely driven by protein destabilisation in the p.Missense Damaging group.

**Figure 2:**
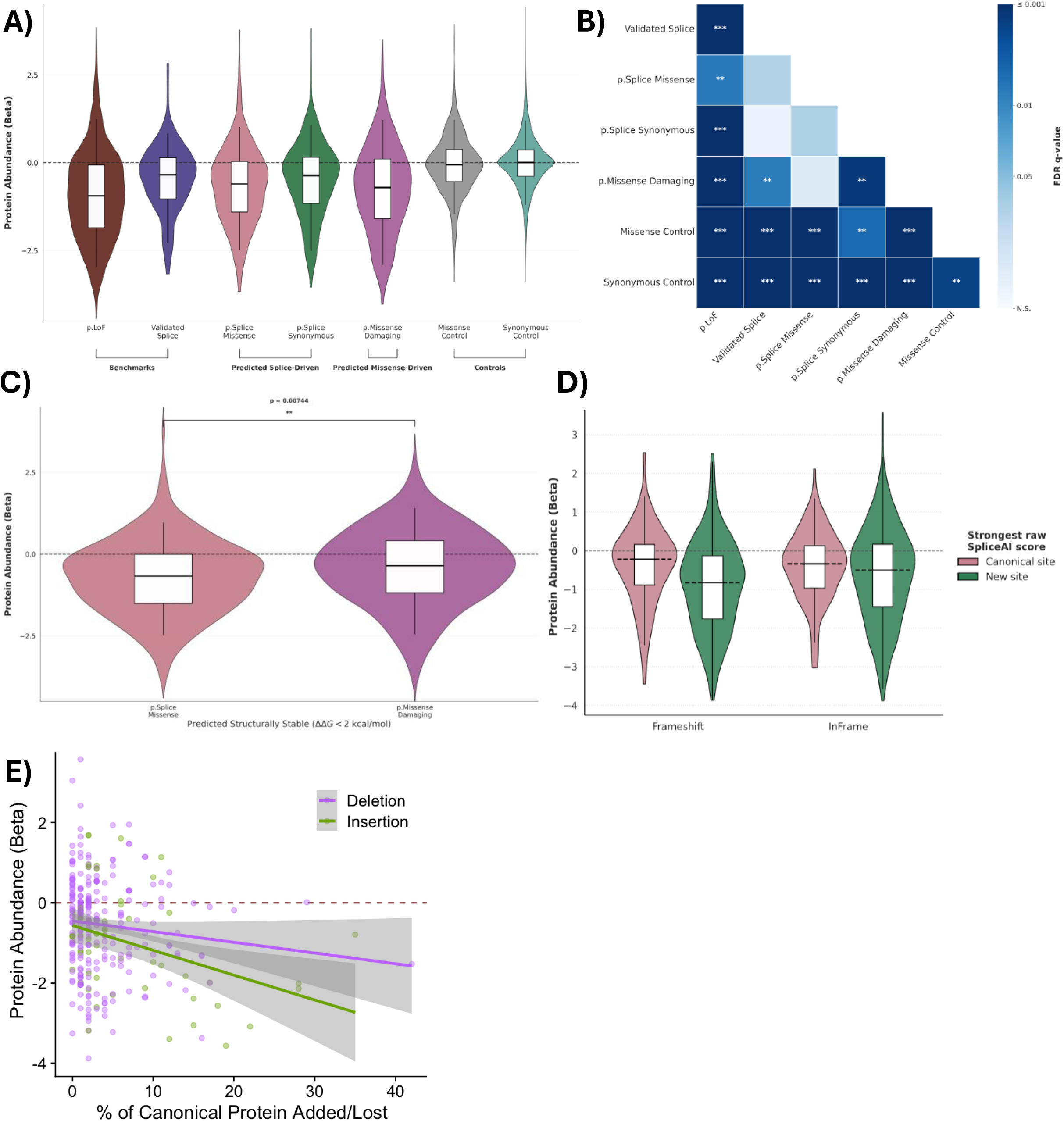
Proteomic validation of the loss-of-function effect in predicted missense spliceogenic variants. (A) Distribution of variant-protein association effect sizes (β) from Olink plasma protein abundance data, stratified across variant groups. Variants are stratified into seven functional categories across four groups: 1) Benchmarks: p.LoF (high-confidence LOFTEE, VEP high-impact, CADD score ≥ 28.1) and Validated Splice (experimentally confirmed in SpliceVarDB). 2) Predicted Splice-Driven: p.Splice Missense (REVEL < 0.5, AlphaMissense < 0.34, SpliceAI Δ ≥ 0.8), and p.Splice Synonymous (SpliceAI Δ ≥ 0.8). 3) Predicted Missense-Driven: p.Missense Damaging (REVEL ≥ 0.932, AlphaMissense ≥ .564, SpliceAI Δ ≤ 0.01). 4) Controls: Missense Control (SpliceAI Δ ≤ 0.01, REVEL < 0.5, AlphaMissense < 0.34), and Synonymous Control (CADD ≤ 0.15, and SpliceAI Δ ≤ 0.01). (B) Pairwise comparisons of Olink protein abundance effect sizes. Heatmap displaying the statistical significance of differences in Olink variant-protein association effect sizes (β) between all variant groups. Significance was determined by a permutation test, with final p-values adjusted for multiple testing via FDR correction. Asterisks denote adjusted q-values: * reflects q-values < 0.05, ** reflects q-values < 0.01, *** reflects q-values < 0.001. (C) Protein abundance for variants predicted to be structurally stable. Comparison of Olink variant-protein association effect sizes (β) for p.Splice Missense and p.Missense Damaging variants subsetted for predicted structural stability (FoldX ΔΔG < 2 kcal/mol). P-value determined by a permutation test. (D) Protein abundance stratified by predicted frame impact and splice junction dominant SpliceAI score. Olink variant-protein effect sizes (β) for missense variants grouped by predicted translational outcome (Frameshift vs In-Frame) based on the primary SpliceAI delta position. Variants are stratified by their greater raw alternate SpliceAI score: canonical splice site (red) or the new site at the delta position (blue). Black dashed lines show group medians. (E) Impact of in-frame alteration length on protein abundance. Scatter plot and linear regression of protein abundance effect sizes (β) for predicted in-frame missense variants against the percentage of canonical protein sequence added (insertion, green) or lost (deletion, purple). Solid lines represent linear regression fits with 95% confidence intervals (shaded regions).

### Splicing-Driven Protein Depletion Occurs Independently of Structural Destabilisation

To investigate whether missense variants cause depletion through destabilisation of the protein, we next controlled for predicted thermodynamic stability by restricting both the HC-LMSVs and p.Missense Damaging groups to variants predicted to maintain their protein’s structural stability (ΔΔG < 2 kcal/mol via FoldX). HC-LMSVs reduced their plasma protein depletion profile further (β_median_ = -0.67, **Figure 2C**), consistent with further enrichment for variants that cause LoF through aberrant splicing. Conversely, removing these structurally destabilising variants markedly reduced the depletion profile of the p.Missense Damaging group, shifting its median abundance effect size from β_median_ = -0.71 to β_median_ = -0.35, consistent with protein destabilisation being the main mechanism of depletion in this group.

There was a significant difference in plasma protein abundance between the HC-LMSVs and structure-maintaining p.Missense Damaging variants (p = 7.4 × 10^-3^), further supporting aberrant splicing – not protein-level effects – as the dominant effect in the MSV group.

### Predicted Translational Frame Disruptions and Novel Splice Site Creation Drives Protein Depletion

We next evaluated whether SpliceAI’s predicted translational frame outcome affects the severity of protein depletion among spliceogenic variants (SpliceAIΔ ≥ 0.2; **Figure 2D**). Variants were grouped by predicted translational outcome (frameshift vs in-frame) and splice site hierarchy, defined by whether the novel cryptic site yielded a higher raw SpliceAI score than the canonical site. We first examined each factor independently across all spliceogenic variants. Variants where a novel cryptic site had a larger raw SpliceAI score than the canonical site demonstrated significantly lower cognate protein abundance than those where the canonical site remained dominant (β_median_ = -0.74, N = 810 versus β_median_ = -0.26, N = 235, Mann-Whitney U p = 2.75 × 10^-7^; **Supplementary Figure S5**). Independently, predicted frameshift alterations induced greater protein depletion than in-frame alterations (β_median_ = -0.71, N = 625 versus β_median_ = -0.45, N = 420, p = 1.23 × 10^-3^).

To determine whether splice site hierarchy and reading frame interact, we stratified variants across both categories. This revealed that the severe protein depletion observed in the novel cryptic site group is heavily driven by frameshifting events. Among predicted frameshifts, variants with a dominant novel site showed marked protein depletion (β_median_ = -0.83; **Figure 2D**) compared to those retaining canonical dominance (β_median_ = -0.23, Dunn’s post-hoc p_adj_ = 8.29 × 10^−6^). Furthermore, within the novel-site dominant group, frameshifts caused significantly greater depletion than in-frame alterations (β_median_ = -0.83 vs β_median_ = -0.5; p_adj_ = 3.7 × 10^-3^). In contrast, for predicted in-frame alterations, splice site dominance had no significant effect on protein abundance (β_median_ = -0.34, p_adj_ = 0.107). Together, these findings demonstrate that raw SpliceAI dominance at novel sites effectively captures disruption at the protein level, with out-of-frame novel sites driving the most severe loss-of-function-like effects, likely resulting in NMD.

### In-Frame Splice Insertions Drive Length-Dependent Protein Depletion

We next investigated whether the magnitude of protein depletion among predicted in-frame spliceogenic variants (using variants from the aforementioned in-frame novel site group) was determined by the extent of the structural disruption to the protein. Stratifying variants by whether the predicted in-frame alteration caused an insertion or deletion and assessing cognate protein abundance effect sizes against the percentage of canonical protein sequence added or lost revealed a divergence between event types (**Figure 2E**).

For in-frame insertions, linear regression showed a statistically significant length-dependent negative correlation (β_intercept_ = -0.57, R^2^ = 0.14, p = 4.58 × 10^-3^), with plasma protein abundance declining by 0.062 standard deviations (β) for every additional 1% of sequence inserted. In contrast, in-frame deletions exhibited a baseline reduction in protein abundance (β = -0.45) but no statistically significant length-dependent relationship (β_slope_ = -0.027, R^2^ = 0.012, p = 0.089). These findings indicate that while in-frame insertions trigger progressive, length-dependent depletion, likely driven by increasingly severe structural disruption, in-frame deletions induce a baseline protein depletion regardless of length. This may suggest that in-frame deletion impact may be determined more by the specific functional or structural elements removed than by length of sequence lost alone.

### Missense Spliceogenic Variants in Intrinsically Disordered Regions Are Severely Discordant With Protein-Centric Predictors

To evaluate whether structural constraint influences the detection of spliceogenicity, we mapped AlphaFold predicted local distance difference test (pLDDT) scores across UKB MANE Select missense variants (N = 3,154,276 unique residues). Intrinsically disordered regions (IDRs), defined by low structural confidence (pLDDT < 50), comprised 5.92% of evaluated UKB residues. Missense variants within IDRs displayed a slightly lower baseline frequency of predicted splice disruption (SpliceAIΔ ≥ 0.2) compared to structured regions (2.41% vs 3.09%; OR = 0.78, p = 1.36 × 10^-95^; **Supplementary Figure S6**). Restricting analysis to the central 70% to control for potential terminal positioning bias generated through enrichment of reduced splice disruption and structural regions, yielded a virtually identical result (OR = 0.79), confirming that this baseline difference is not an artefact of terminal enrichment. However, when a splice disruption is predicted to occur within an IDR (SpliceAIΔ ≥ 0.2), protein-centric pathogenicity predictors often fail to predict any potential disruption. Among MSVs, 85.2% occurring in IDRs were LMSVs, compared to 53.5% in structured domains (OR = 5, p = 0). Therefore, while predicted splice disruption is marginally less frequent in IDRs, MSVs in disordered regions are five times more likely to differ in predicted classification compared to protein-centric predictors.

### Patient Transcriptomics Validate Predicted Cryptic Splicing of ClinVar VUSs

To evaluate the broader clinical landscape of predicted MSVs, we analysed the distribution of SpliceAIΔ scores among 1,947,273 missense variants in ClinVar, which we stratified by their reported clinical significance (**Supplementary Table S4**). Notably, at the sensitive threshold (SpliceAIΔ ≥ 0.2), the proportion of predicted MSV-VUSs (2.46%) closely mirrors the UKB population background (2.71%). However, at the high-precision threshold (SpliceAIΔ ≥ 0.8), the proportion of MSV-VUSs falls significantly to 0.06%. This decline represents a five-fold depletion relative to the UKB baseline (0.31%) and mirrors the proportion we observed in the Benign/Likely Benign tier (0.07%). Evaluating the composition of these predicted spliceogenic variants (Δ ≥ 0.2) across the tiers revealed that while high-confidence predicted MSVs (Δ ≥ 0.8) constitute 2.4% of the VUSs, they account for 38.1% of all predicted MSVs within the P/LP tier, representing a nearly 16-fold enrichment of high-confidence scores among P/LP variants relative to VUSs.

To systematically evaluate LMSVs currently reported as VUSs in ClinVar, we integrated the ClinVar VUS dataset with the Genomics England 100kGP RNA-seq data. Filtering 537,127 ClinVar VUSs in developmental disorder-associated genes from PanelApp resulted in 6,709 LMSVs, of which 1,311 were tiered within the 100kGP cohort. Cross-referencing these variants against participants with transcriptomic data yielded 129 variants across 169 individuals. We excluded 67 (51.9%) due to lack of evidence upon which to determine a splice effect due to a combination of either low/no coverage of the target loci, the variant not being expressed, or the variant’s reads not spanning the splice junction (**Additional Files**).

We identified 15 missense VUSs out of 62 (24.2%) with robust transcriptomic evidence to support a splice disruption event (**Additional Files**). This included a deep exonic missense variant in *PREPL* (NM_001171613.2:c.1216C>T) in a single heterozygous carrier (**Figure 3**). Biallelic loss-of-function of this gene is associated with autosomal recessive congenital hypotonia-cystinuria syndrome (MIM: 616224). Consistent with SpliceAI predictions (**Figure 3A**), haplotype-resolved read alignment demonstrated that the variant causes aberrant splicing; multiple reads confirmed that the variant activates an exonic cryptic splice site located 2-bp upstream of the variant (**Figure 3B**). Across 69 delivery batch-matched controls, we observed zero usage of this cryptic splice site (Fisher’s Exact Test p = 2.17 × 10^-8^, Odds Ratio [OR] = ∞), confirming its unique presence in the proband. This splicing event induces an inframe 48-bp exonic deletion, resulting in the loss of 16 highly conserved amino acids comprising a β-hairpin within a central β-sheet (**Figure 3C**).

**Figure 3:**
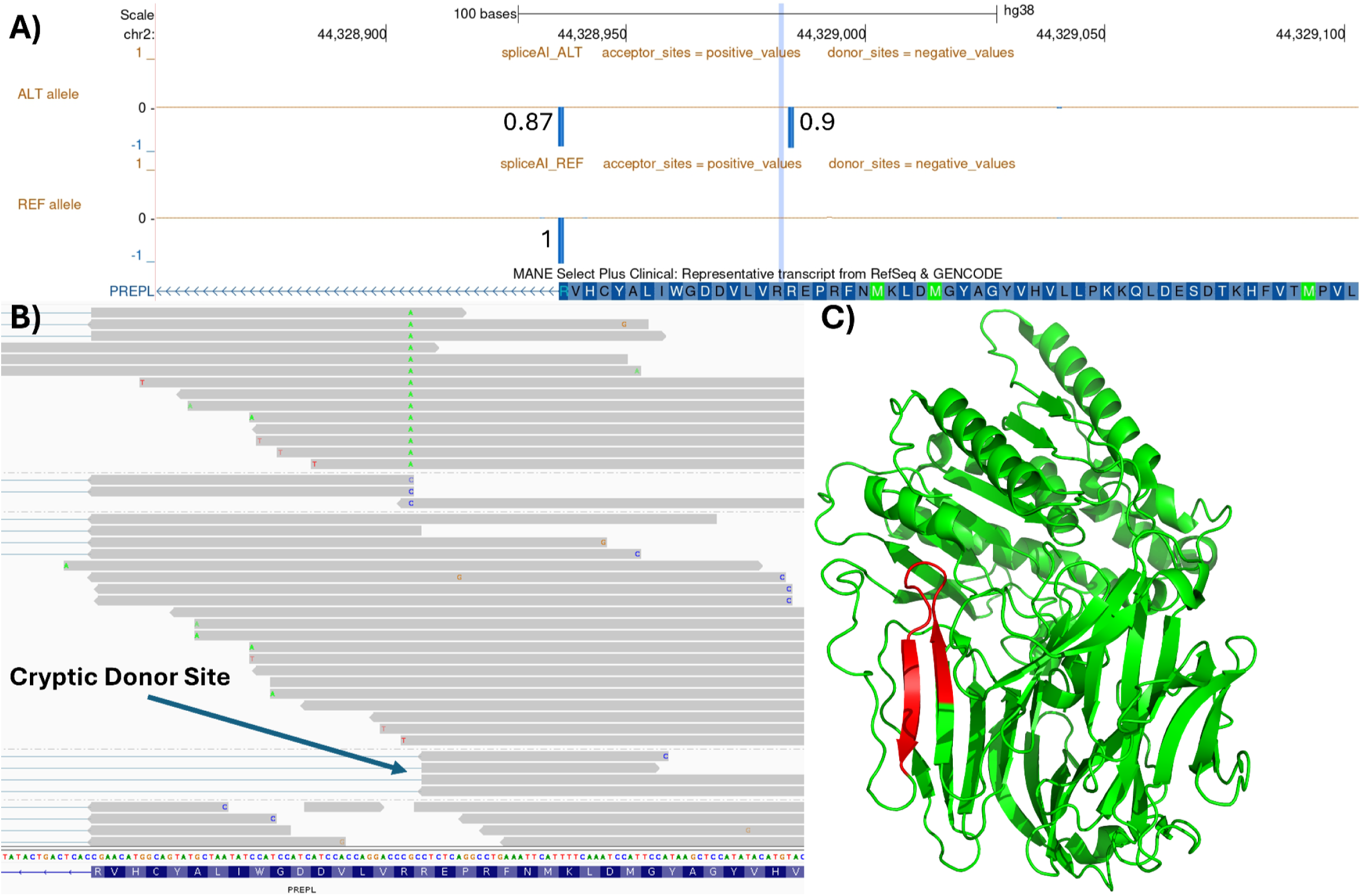
Functional and predicted structural validation of a cryptic spliceogenic missense *PREPL* variant. (A) SpliceAI prediction of a cryptic donor gain. UCSC Genome Browser track displaying the *PREPL* NM_001171613.2:c.1216C>T p.(Arg406Trp) variant locus. The light blue highlight represents the variant’s position. The SpliceAI raw alternate scores are found on the top track; reference scores are found on the bottom track. (B) Transcriptomic validation of aberrant splicing in the 100kGP. IGV screenshot demonstrating the presence of the predicted cryptic splice junction in a 100kGP carrier. Use of this cryptic donor site was not seen in 69 delivery-matched 100kGP control individuals (Fisher’s Exact p = 2.2 × 10^-8^, OR = ∞). (C) Predicted structural impact of aberrant splicing on the PREPL protein (PDB7OBM). Protein structure of PREPL with the resultant 16 amino acid in-frame deletion highlighted in red.

### Retrospective Evaluation of Solved Cohorts Reveals Pathogenic Missense Spliceogenic Variants causing Haploinsufficiency

To evaluate if MSVs have been previously identified as diagnostic, we performed a retrospective analysis of solved cases across the DDD study and the 100kGP cohort. In the DDD study, we limited our analysis to previously identified pathogenic/likely pathogenic missense variants publicly available in DECIPHER^25^. We identified 73 publicly accessible, clinically assessed pathogenic/likely pathogenic missense variants with SpliceAIΔ ≥ 0.2 in 87 DDD probands, of which eight variants in nine probands were LMSVs in which aberrant splicing is the most likely pathomechanism (**Additional Files**). Notably, this subset included a *de novo* missense variant in *ARID1B* (NM_001374828.1:c.2581G>A), an intrinsically disordered protein in which amino acid changes are generally tolerated but where haploinsufficiency causes Coffin-Siris syndrome (MIM: 135900).

In the 100kGP, we extended our retrospective analysis to all tiered variants from 9,046 solved probands within the Genomics England 100kGP, focusing strictly on LMSVs. This isolated 9,225 tiered LMSVs across the cohort, of which six were classified as pathogenic (**Additional Files**). Five of the six LMSVs have been functionally validated as splice disrupting, as evidenced in their ClinVar records.

### Targeted Validation of Pathogenic Missense Spliceogenic Variants in a Clinical Cohort Supports Pathomechanism

We characterised 14 missense VUS that were flagged by the NHS Genomic Medicine Service as likely causes of disease through loss-of-function caused by aberrant splicing, all possessing a SpliceAIΔ ≥ 0.2 (**Table 1**). This cohort serves as a real-world example of the potential diagnostic outputs yielded when embedding splice prediction tools into missense interpretation. Together, the combination of *in-silico* splice predictions with RNA-sequencing evidence demonstrating aberrant splicing, and variant prioritisation by the referring clinical genetics service supports considering all 14 variants as pathogenic or likely pathogenic.

**Table 1:** Fourteen experimentally validated missense spliceogenic variants prioritised by the NHS Genomic Medicine Service. AR: Autosomal Recessive, AD: Autosomal Dominant, SCD: Splicing and Disease Study

| Variant | Gene | Mode of Inheritance | Genotype | SpliceAIΔ | REVEL | AlphaMissense | Experimental Evidence | Patient Phenotype | MIM Number | Cohort |
| --- | --- | --- | --- | --- | --- | --- | --- | --- | --- | --- |
| NM_005807.6:c.3991G>A, p.(Gly1331Ser) | PRG4 | AR | Heterozygous | 0.66 | 0.180 | 0.2981 | Novel donor site (98bp loss) | Polyarthropathy | #208250 | S&D |
| NM_003384.3:c.1160G>T p.(Arg387Leu) | VRK1 | AR | Homozygous | 0.60 | 0.068 | 0.2278 | Exon Skipping | Progressive Gait Ataxia | #607596 | S&D |
| NM_004247.4:c.1393A>G p.(Met465Val) | EFTUD2 | AD | Heterozygous | 0.99 | 0.453 | 0.2244 | Novel Donor Site r.1393_1411del, p.(Ser466AlafsTer3) | Left Kidney Agenesis, Klippel-Feil, Scoliosis, Arachnoid Cyst | %601076 | S&D |
| NM_004238.3:c.4315C>T p.(His1439Tyr) | TRIP12 | AD | Heterozygous | 0.98 | 0.136 | 0.2956 | r.4314_4318del | No Consent to Publish | - | NHS |
| NM_000059.4:c.425G>A p.(Ser142Asn) | BRCA2 | AD | Heterozygous | 0.91 | 0.153 | 0.4471 | Exon 4 Skipping r.316_425del p.(Gly106SerfsTer15) | 3 Generations of Breast Cancer | #612555 | S&D |
| NM_025132.4:c.1835T>A p.(Leu612Gln) | WDR19 | AR | Heterozygous | 0.97 | 0.894 | 0.4404 | Novel donor site | Skeletal Dysplasia | *608151 | S&D |
| NM_000267.3:c.7832A>G, p.(Asp2632Gly) | NF1 | AD | Heterozygous | 0.43 | 0.419 | 0.4781 | Exon Skipping r.7870_7970del, p.Thr(2625Ter) | Neurofibromatosis with Multiple Neuromas | #162200 | S&D |
| NM_000548.5:c.4492A>C p.(Ser1498Arg) | TSC2 | AD | Heterozygous | 0.41 | 0.939 | 0.9837 | Novel donor site r.4240_4493del, p.(Val1414PhefsTer24) | Tuberous Sclerosis | #613254 | S&D |
| NM_001378452.1:c.1712A>G p.(Gln571Arg) | ITPR1 | AD | Heterozygous | 0.55 | 0.921 | 0.994 | Novel Donor Site | Ataxic Cerebral Palsy | #117360 | S&D |
| NM_004612.4:c.574G>C p.(Gly192Arg) | TGFBR1 | AD | Heterozygous | 0.97 | 0.911 | 0.9998 | Exon Skipping r.344_574del | No Consent to Publish | - | NHS |
| NM_000303.3:c.178G>A p.(Val60Met) | PMM2 | AR | Heterozygous | 0.53 | 0.774 | 0.4365 | Intronic Insertion r.178_179ins178+1_178+479, p.(Val60GlyfsTer3) | No Consent to Publish | - | NHS |
| NM_000314.8:c.253G>C p.(Val85Leu) | PTEN | AD | Heterozygous | 0.55 | 0.919 | 0.9937 | Exon Skipping, Novel Donor Site, Cryptic Exon r.210_253del | No Consent to Publish | - | NHS |
| NM_000249.4:c.704A>G p.(Asp235Gly) | MLH1 | AD | Heterozygous | 0.29 | 0.85 | 0.5159 | Exon 9 Skipping | Colon Cancer | #120435 | S&D |
| NM_000138.5:c.3964G>C p.(Asp1322His) | FBN1 | AD | Heterozygous | 0.97 | 0.973 | 0.9971 | r.3839_3964del | No Consent to Publish | - | S&D |

Notably, four variants had consistently low protein-centric pathogenicity scores. This included a homozygous missense variant in *VRK1,* in which biallelic loss-of-function causes autosomal recessive pontocerebellar hypoplasia type 1A (MIM: 607596). The variant (NM_003384.3:c.1160G>T) had a high-confidence splice prediction (SpliceAIΔ = 0.60), yet protein-centric predictors misclassified it as benign (REVEL = 0.068; AlphaMissense = 0.2278). We performed RNA-seq on the affected proband and confirmed that the variant disrupts exon 13’s canonical acceptor site, driving exon skipping and the use of an alternate transcript’s final exon (ENST00000680335.1), along with retention of intron 12. This splice-disruption results in the proband’s progressive gait ataxia phenotype (**Figure 4**). Another validated pathogenic LMSV was a missense variant in *PRG4,* present in compound heterozygosity with a loss-of-function variant, where biallelic loss-of-function variants cause camptodactyly-arthropathy-coxa vara-pericarditis syndrome (as detailed by Masroori et al.^42^; MIM: 208250). The variant (NM_005807.6:c.3991G>A) displayed a high SpliceAI score (SpliceAIΔ = 0.66) despite benign missense pathogenicity scores (REVEL = 0.18; AlphaMissense = 0.2981). We used gel electrophoresis and Sanger sequencing to confirm that this variant disrupts the canonical exon 11 donor site, forcing the use of a cryptic upstream donor site that generates a 97-bp exonic deletion in a proband presenting with polyarthropathy (**Figure 5**).

**Figure 4:**
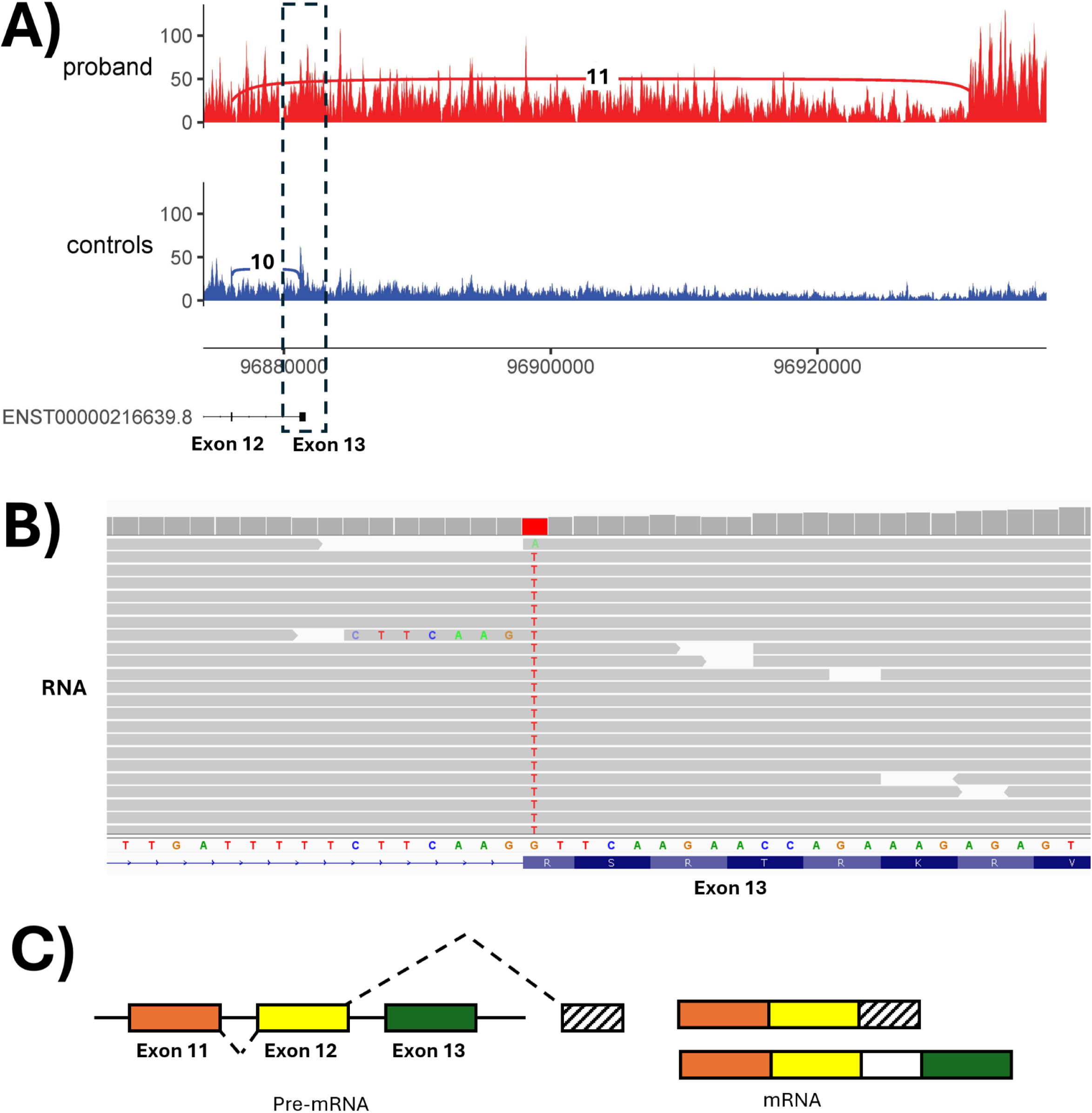
Experimental validation of NM_003384.3(*VRK1*):c.1160G>T. (A) Sashimi plot showing the nature of the splicing disruption in the individual with the NM_003384.3:c.1160G>T variant (red) relative to RNA-seq from ten other individuals without that variant (blue). (B) IGV screenshot of RNA sequencing reads showing the G>T variant within exon 13 and intron retention. (C) Schematic diagram summarising the observed impact(s) on splicing.

**Figure 5:**
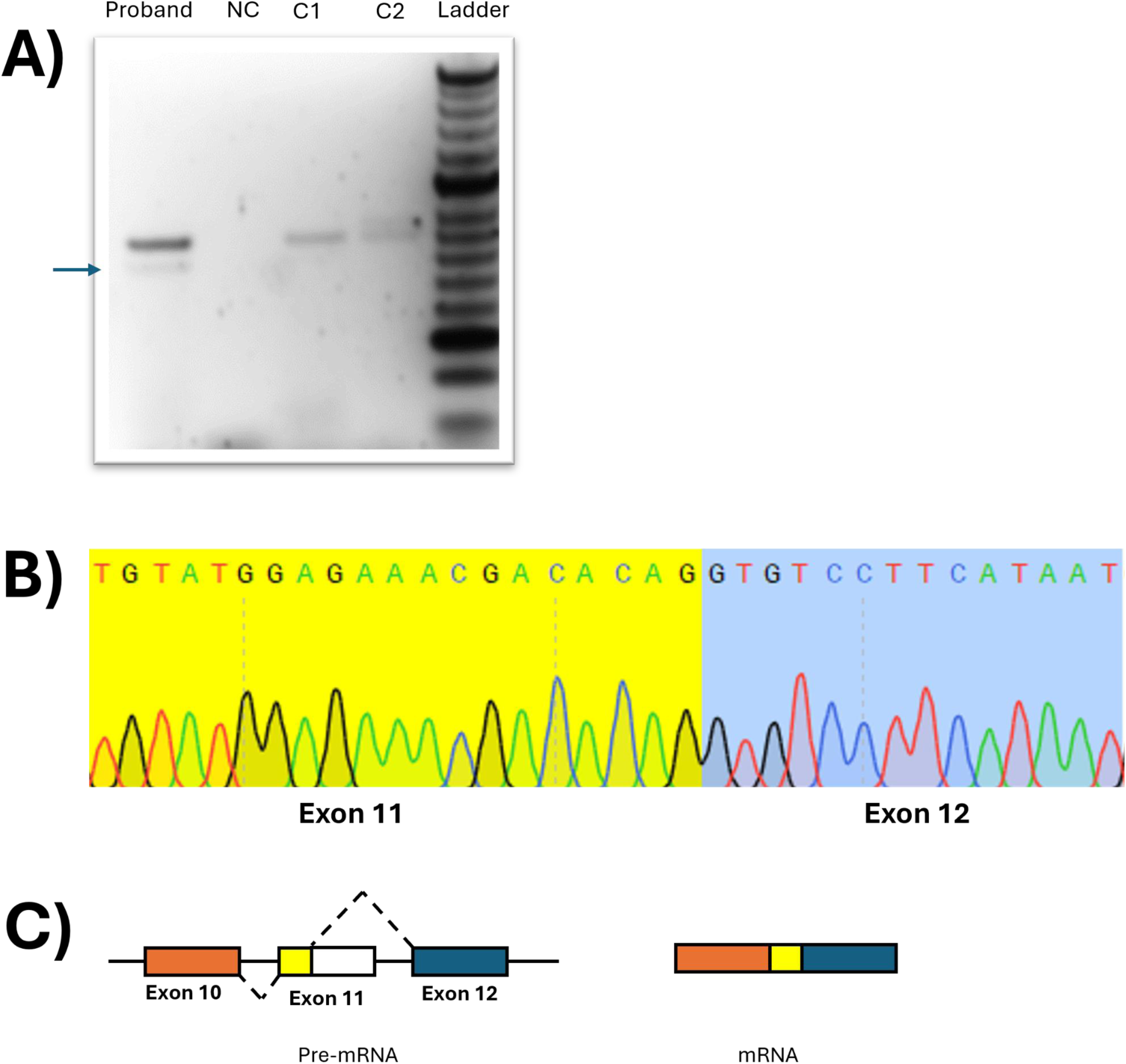
Experimental validation of NM_005807.6(*PRG4*):c.3GG1G>A. (A) Agarose gel electrophoresis of RT-PCR products of NM_005807.6:c.3991G>A variant. A distinct band corresponding to the expected amplicon size after aberrant splicing is observed in the sample lane (arrow indicates representative band). (B) Sanger sequencing chromatogram of the RT-PCR amplicon confirming 97 base pair deletion. (C) Schematic diagram summarising the observed impact on splicing. NC: Negative Control, C1, C2: Controls

Among the remaining 10 variants, two scored below our defined tolerated thresholds for at least one of the two missense tools (REVEL < 0.5 or AlphaMissense < 0.34), three were predicted pathogenic by REVEL and ambiguous by AlphaMissense (0.34 < × < 0.564), and the remaining five were predicted pathogenic by both predictors (**Table 1**). However, because these tools are intrinsically protein-centric, they fail to capture the spliceogenic mechanism, potentially misdirecting downstream functional validation analysis and failing to highlight a relevant pathomechanism.

### Prioritisation Framework Identifies Diagnostic Variants in the Unsolved 100kGP Cohort

To demonstrate the benefits of using splice prediction tools on missense variants to resolve unsolved cases, we assessed tiered homozygous and *de novo* LMSVs in probands lacking a diagnosis in the 100kGP. We identified 1,366 unsolved participants who were homozygous for one of the 9,225 tiered LMSVs and 47 who had *de novo* LMSVs. Restricting to variants in disease-associated genes with the appropriate mode of inheritance resulted in 113 candidate homozygous variants across 270 participants, and zero *de novo* variants. To further prioritise high-confidence diagnostic candidates, we applied stringent population frequency filtering (gnomAD AF < 0.01, < 3 gnomAD homozygotes, 0 unaffected homozygotes across 100kGP and UKB) and manually evaluated participant clinical records with expected gene-disease phenotypes (**Methods**). This filtering pipeline narrowed the candidate list to two diagnostic candidates that strongly match participant phenotypes and warrant further clinical examination.

Our first candidate was a homozygous variant in *DYSF* recently described by *Lord et al.*^24^ in which biallelic loss-of-function variants cause autosomal recessive muscular dystrophy (MIM: 253601). The variant (NM_001130987.2:c.6173G>A) is reported as conflicting in ClinVar (VUS versus Likely Pathogenic, VCV000285133.10), with a SpliceAIΔ of 0.42, a REVEL of 0.256, and an AlphaMissense of 0.0669. We performed a retrospective transcriptomic assessment of this proband, who presents with limb-girdle muscular dystrophy, and confirmed that the variant drives the loss of exon 54’s canonical donor site, triggering the use of a cryptic splice site 143 bp downstream.

Our second candidate was a homozygous variant in *XRCC1*(NM_006297.3:c.1293G>C), a gene in which biallelic loss-of-function variants cause autosomal recessive spinocerebellar ataxia (MIM: 617633). The variant was identified in two unsolved, unrelated probands of South Asian descent presenting with hereditary ataxia. To date, only four patients harbouring this variant have been published in the literature^43,44^, all of South Asian ancestry.

Homozygosity mapping in two Pakistani probands not known to be related identified a shared region of homozygosity on chromosome 19 encompassing *XRCC1*, and the variant has subsequently been described as a South Asian founder allele^44^. Identifying two additional South Asian probands expands the reported cohort to six cases and provides further evidence for this founder variant. Although we currently lack proband transcriptomic data for validation, the variant demonstrates strong evidence of being an MSV (SpliceAIΔ = 0.95; REVEL = 0.052; AlphaMissense = 0.2415), and there is a strong clinical fit with the patient phenotypes.

## Discussion

This study provides the first genome-wide estimate of the proportion of missense spliceogenic variants (MSVs), and uses a variety of transcriptomic, proteomic and clinical evidence to support aberrant splicing. We estimate that around 2.7% of missense variants may disrupt splicing, sharing a baseline spliceogenic burden with synonymous variants that extends at least 19 bases into exons. We show that MSVs cause a significant depletion in circulating protein levels, even when the amino acid substitution is predicted to be tolerated. Furthermore, we provide 32 examples of rare disease probands across the DDD Study, 100kGP and NHS GMS in which a pathogenic missense variant likely explains their disease via aberrant splicing (**Supplementary Table S5**).

These findings challenge the widespread practice of interpreting exonic variants primarily according to predicted coding consequences. Existing variant classification frameworks^9,11^ primarily recommend evaluating missense variants based on protein position (e.g. proximity to binding sites and known hotspots) and using protein-centric predictors (such as AlphaMissense and REVEL), which prioritise protein structural effects and evolutionary conservation. However, discovering that 82% of predicted MSVs were classified as benign or neutral by at least one of these tools (and 57% by both) highlights an important limitation of protein-centric interpretation alone. This observation translated into our clinical work, with 77% (n = 27/35) of experimentally supported MSVs also exhibiting this discordance, including 69% (n = 24/35) across both protein-centric tools. Notably, the same discordance was also observed in deep exonic missense variants located up to 19 bp from canonical splice junctions, which fall outside the canonical splice region yet retain an elevated predicted spliceogenic burden. Together, these findings suggest that incorporating position-independent splice prediction alongside protein-centric assessment could reduce the likelihood that MSVs are overlooked during variant prioritisation and interpretation.

Evaluating missense variants at the transcript-level may also aid clinical interpretation by supporting a loss-of-function mechanism (e.g. PVS1-equivalent evidence^45^). This is particularly relevant in proteins where amino acid substitutions do not generally cause disease but where loss-of-function is an established pathomechanism. This is true within intrinsically disordered regions (IDRs), which comprise approximately 35%^46^ of residues across the human proteome. As IDRs lack fixed tertiary structure and generally exhibit lower evolutionary conservation, protein-centric tools frequently assign benign or ambiguous pathogenicity scores to IDR missense variants, contributing to 85.7%^47^ of ClinVar missense variants in IDRs being classified as VUSs. Crucially, because mRNA splicing occurs independently of downstream protein folding, missense splice-disruption is predicted in IDRs at rates comparable to structured domains (2.41% vs 3.09%). Yet, when splice disruption is predicted to occur within an IDR, protein-centric tools fail to predict such events in 85.2% of cases (OR = 5). Therefore, transcript-level interpretation may rescue hidden loss-of-function variants in regions traditionally considered tolerant to missense variation, exemplified by our identification of a diagnostic missense variant in *ARID1B* within the DDD cohort predicted to be spliceogenic.

We leveraged the UKB’s proteomic data to confirm transcript-mediated protein depletion and discern the effect from structural destabilisation leading to protein depletion via misfolding.

By applying a stringent predictive thermodynamic filter (ΔΔG < 2 kcal/mol) via FoldX to identify structurally stable proteins, we isolated two sub-groups enriched for variants likely acting through transcript-level versus protein misfolding mechanisms. Following filtering, the predicted MSV group maintained a loss-of-function depletion profile, whereas the predicted damaging missense group halved its depletion profile compared with destabilising variants. This divergence is consistent with transcript-level depletion following splice disruption (likely via NMD), rather than protein misfolding, as the predominant mechanism for the observed reduction in protein abundance.

Our findings further highlight a key limitation of using the standard SpliceAI output, in that the traditional delta scores (Δ) quantify relative changes in splicing probability, but fails to evaluate whether the new cryptic site actually outscores the canonical site to become the high probability splice junction. By integrating splice site hierarchy – comparing raw SpliceAI scores between novel and canonical sites – we show that severe protein depletion is heavily concentrated in novel-site-dominant events, particularly predicted frameshift events. In contrast, variants where the canonical site retains dominance, or those predicted to cause in-frame alterations that bypass NMD, exhibit milder or variable protein depletion likely driven by structural constraints. This highlights how relying solely on standard delta scores without assessing the raw scores risks over-interpreting splice predictions in which the canonical site remains dominant.

While our framework effectively captures MSVs at scale, several cohort-specific, technical, and biological constraints warrant consideration. Firstly, the UK Biobank reflects a healthy population cohort (recruited ages 40-69) of predominantly EUR-like ancestry. This demographic introduces survivorship bias through the depletion of severe, early-onset disease-causing variants, so our prevalence estimates likely represent an underestimate that may not generalise across diverse global populations. Secondly, proteomic validation was restricted to circulating plasma proteins measured via Olink PEA assays, which fails to capture intracellular or non-secreted protein targets along with tissue-specific effects, and remains susceptible to epitope-disrupting artefacts that can mimic protein depletion. Thirdly, transcriptomic validation relied on whole-blood RNA sequencing, leaving tissue-specific splicing events unexamined and low-expression loci vulnerable to zero read coverage.

Finally, functional validation of splice disruption is intrinsically obscured by active NMD, which rapidly degrades transcripts containing premature termination codons and reduces reads spanning both the variant and a splice junction required for detection, alongside short-read sequencing constraints that hinder phasing deep exonic missense variants with spliceogenic effects.

While we focused exclusively on the widely-used SpliceAI in this study, which provides a strong foundation for splice prediction, complementary tools like Pangolin^15^ (provides Percent Spliced In [**Ψ**] values), AbSplice2^48^ (tissue- and development-specific), MaxEntScan^16–18^ (optimal performance with SpliceAI) and AlphaGenome^49^ may further improve interpretation by capturing alternative aspects of splicing. Finally, while the UKB SpliceAI outputs underwent reannotation (to increase the SpliceAI window from 50bp to 500bp) and rigorous curation, the clinical cohorts relied on pre-computed scores. As documented by Martin-Geary et al.^30^, the pre-computed SpliceAI carry limitations including improper variant masking, a limited 50 base pair window size, and an outdated annotation file (e.g. GENCODE V24lift37).

Our identification of an unsolved proband with limb-girdle muscular dystrophy harbouring a homozygous MSV in *DYSF* illustrates the clinical value of incorporating transcript-centric models early in missense variant interpretation. Although this participant’s genomic data had been available since 2017, the absence of systematic splice assessments for missense variants during routine tiering likely contributed to a prolonged diagnostic delay. This case study demonstrates how MSVs may be overlooked when variant interpretation and prioritisation relies primarily on protein-centric evidence. Our work supports incorporating position-independent splice predictions into the computational evidence used for missense variant prioritisation and interpretation within variant classification frameworks^9,11^. Such an approach would complement existing protein-centric interpretation and could improve the prioritisation of clinically relevant MSVs, helping to reduce the diagnostic odyssey for patients with rare disease.

## Data Availability

Custom Python scripts used for splice-junction annotation, transcriptomic screening, and downstream analyses are available at https://github.com/MRintari/Missense-Spliceogenic-Variant-Analysis. The modified SpliceAI code is available at https://github.com/chundruv/SpliceAImod.git.
Data from the National Genomic Research Library (NGRL) used in this research are available within the secure Genomics England Research Environment. Access to NGRL data is restricted to adhere to consent requirements and protect participant privacy. Data used in this research include primary NGRL datasets such as Rare Disease Tiering Data and Clinical Exit Questionnaires accessed via LabKey. Individual-level candidate variant records and clinical details were reviewed using the Participant Explorer and the Interactive Variant Analysis Case Portal and Variant Browser. Annotation scores for SpliceAI and AlphaMissense were accessed through the environment public data resources directory, while REVEL scores were obtained directly through participant Variant Effect Predictor annotation files. Access to NGRL data is provided to approved researchers who are members of the Genomics England Research Network, subject to institutional access agreements and research project approval under participant-led governance. For more information on data access, visit: https://www.genomicsengland.co.uk/research

## Data and Code Availability

Custom Python scripts used for splice-junction annotation, transcriptomic screening, and downstream analyses are available at https://github.com/MRintari/Missense-Spliceogenic-Variant-Analysis. The modified SpliceAI code is available at https://github.com/chundruv/SpliceAImod.git.

Data from the National Genomic Research Library (NGRL) used in this research are available within the secure Genomics England Research Environment. Access to NGRL data is restricted to adhere to consent requirements and protect participant privacy. Data used in this research include primary NGRL datasets such as Rare Disease Tiering Data and Clinical Exit Questionnaires accessed via LabKey. Individual-level candidate variant records and clinical details were reviewed using the Participant Explorer and the Interactive Variant Analysis’ Case Portal and Variant Browser. Annotation scores for SpliceAI and AlphaMissense were accessed through the environment’s public data resources directory, while REVEL scores were obtained directly through participant Variant Effect Predictor annotation files. Access to NGRL data is provided to approved researchers who are members of the Genomics England Research Network, subject to institutional access agreements and research project approval under participant-led governance. For more information on data access, visit: https://www.genomicsengland.co.uk/research

## Acknowledgments

This research has been conducted using the UK Biobank Resource under Application Number 103356. This research was funded in whole or in part by the Wellcome Trust [226083/Z/22/Z] and was supported by the National Institute for Health and Care Research Exeter Biomedical Research Centre. The views expressed are those of the author(s) and not necessarily those of the NIHR or the Department of Health and Social Care. For the purpose of open access, the author has applied a CC-BY public copyright licence to any author accepted manuscript version arising from this submission.

We thank the patients and families involved in the NHS-GMS, 100kGP and DDD Study. Ethical approval for the 100,000 Genomes Project was initially provided by Cambridge South REC (14/EE/1112) but recently an updated version was approved by East of England - Cambridge Central Research Ethics Committee (25/EE/0125). We gratefully acknowledge the participants of the National Genomic Research Library (NGRL), whose contributions made this research possible. Secure access to the NGRL under project ID 349 was provided by Genomics England, which delivers the NGRL in partnership with NHS England, and is wholly owned by the UK Department of Health and Social Care. The NGRL contains participants’ health data collected by the NHS as part of their care, along with samples and data from their participation in research, for which fully informed consent has been obtained. This includes genomic and clinical data provided through the NHS Genomic Medicine Service, as well as data obtained through research studies, including the 100,000 Genomes Project and the Generation Study, both of which are delivered in partnership with the NHS, and from other research cohorts involving external collaborators.

The authors thank all patients, families, and clinicians involved in the University of Southampton’s Splicing and Disease study (IRAS 49685).

## Author Contributions

M.R.R, C.F.W., and V.K.C. conducted conceptualisation of this study. M.R.R., V.K.C., C.F.W., C.J.O., and D.B. conducted data curation. M.R.R. and V.K.C. carried out the formal analysis. C.F.W. and D.B. acquired funding for this project. M.R.R., V.K.C., C.J.O., J.L., and A.T.P. conducted the investigation within this study. M.R.R., C.F.W., V.K.C., C.J.O., J.L, A.T.P., and D.B. contributed to method development. M.R.R., C.F.W., and V.K.C. were responsible for project administration. C.F.W., C.J.O., and D.B. provided resources used in this study. M.R.R. and V.K.C. were responsible for the development of software used in this study. C.F.W. and V.K.C. conducted supervision throughout the study. M.R.R. and V.K.C. conducted validation of results throughout the study. M.R.R., V.K.C., and C.J.O. conducted visualisation in this study. M.R.R., V.K.C., and C.J.O wrote the manuscript, with input and revisions provided by C.F.W., J.L., A.T.P, and D.B. All authors approved the final manuscript.

## Web Resources

### Supplemental Information

**Document S1. Figures S1-S4**

**Data S1. Tables S1-S5**

**Data S2. Additional Tables Sheets 1-3**

## Declaration of Generative AI and AI-Assisted Technologies in the Writing Process

During the preparation of this work the author(s) used Google Gemini in order to improve the readability and language of the text. After using this tool, the author(s) reviewed and edited the content as needed and takes full responsibility for the content of the publication.

## References

1. Palafox, M.F., Boatner, L., Wilde, B.R., Christofk, H., Backus, K.M., and Arboleda, V.A. (2025). Prioritizing disease-associated missense variants with chemoproteomic-detected amino acids. Am. J. Hum. Genet. 112, 1649–1663. 10.1016/j.ajhg.2025.04.017.

2. Stenson, P.D., Mort, M., Ball, E.V., Shaw, K., Phillips, A.D., and Cooper, D.N. (2014). The Human Gene Mutation Database: building a comprehensive mutation repository for clinical and molecular genetics, diagnostic testing and personalized genomic medicine. Hum. Genet. 133, 1–9. 10.1007/s00439-013-1358-4.

3. Liu, H.-X., Cartegni, L., Zhang, M.Q., and Krainer, A.R. (2001). A mechanism for exon skipping caused by nonsense or missense mutations in BRCA1 and other genes. Nat. Genet. 27, 55–58. 10.1038/83762.

4. Mort, M., Sterne-Weiler, T., Li, B., Ball, E.V., Cooper, D.N., Radivojac, P., Sanford, J.R., and Mooney, S.D. (2014). MutPred Splice: machine learning-based prediction of exonic variants that disrupt splicing. Genome Biol. 15, R19. 10.1186/gb-2014-15-1-r19.

5. Krawczak, M., Thomas, N.S.T., Hundrieser, B., Mort, M., Wittig, M., Hampe, J., and Cooper, D.N. (2007). Single base-pair substitutions in exon–intron junctions of human genes: nature, distribution, and consequences for mRNA splicing. Hum. Mutat. 28, 150–158. 10.1002/humu.20400.

6. Lim, K.H., Ferraris, L., Filloux, M.E., Raphael, B.J., and Fairbrother, W.G. (2011). Using positional distribution to identify splicing elements and predict pre-mRNA processing defects in human genes. Proc. Natl. Acad. Sci. 108, 11093–11098. 10.1073/pnas.1101135108.

7. Sterne-Weiler, T., Howard, J., Mort, M., Cooper, D.N., and Sanford, J.R. (2011). Loss of exon identity is a common mechanism of human inherited disease. Genome Res. 21, 1563–1571. 10.1101/gr.118638.110.

8. Thompson, B.A., Walters, R., Parsons, M.T., Dumenil, T., Drost, M., Tiersma, Y., Lindor, N.M., Tavtigian, S.V., de Wind, N., Spurdle, A.B., et al. (2020). Contribution of mRNA Splicing to Mismatch Repair Gene Sequence Variant Interpretation. Front. Genet. 11. 10.3389/fgene.2020.00798.

9. Richards, S., Aziz, N., Bale, S., Bick, D., Das, S., Gastier-Foster, J., Grody, W.W., Hegde, M., Lyon, E., Spector, E., et al. (2015). Standards and guidelines for the interpretation of sequence variants: a joint consensus recommendation of the American College of Medical Genetics and Genomics and the Association for Molecular Pathology. Genet. Med. 17, 405–424. 10.1038/gim.2015.30.

10. Chen, E., Facio, F.M., Aradhya, K.W., Rojahn, S., Hatchell, K.E., Aguilar, S., Ouyang, K., Saitta, S., Hanson-Kwan, A.K., Capurro, N.N., et al. (2023). Rates and Classification of Variants of Uncertain Significance in Hereditary Disease Genetic Testing. JAMA Netw. Open 6, e2339571. 10.1001/jamanetworkopen.2023.39571.

11. Durkie, M., Cassidy, E.-J., Berry, I., Owens, M., Turnbull, C., Taylor, R.W., Deans, Z.C., Ellard, S., and Baple, E.L. ACGS Best Practice Guidelines for Variant Classification in Rare Disease 2024.

12. Cheng, J., Novati, G., Pan, J., Bycroft, C., Žemgulytė, A., Applebaum, T., Pritzel, A., Wong, L.H., Zielinski, M., Sargeant, T., et al. (2023). Accurate proteome-wide missense variant effect prediction with AlphaMissense. Science 381, eadg7492. 10.1126/science.adg7492.

13. Ioannidis, N.M., Rothstein, J.H., Pejaver, V., Middha, S., McDonnell, S.K., Baheti, S., Musolf, A., Li, Q., Holzinger, E., Karyadi, D., et al. (2016). REVEL: An Ensemble Method for Predicting the Pathogenicity of Rare Missense Variants. Am. J. Hum. Genet. 99, 877–885. 10.1016/j.ajhg.2016.08.016.

14. Jaganathan, K., Kyriazopoulou Panagiotopoulou, S., McRae, J.F., Darbandi, S.F., Knowles, D., Li, Y.I., Kosmicki, J.A., Arbelaez, J., Cui, W., Schwartz, G.B., et al. (2019). Predicting Splicing from Primary Sequence with Deep Learning. Cell 176, 535–548.e24. 10.1016/j.cell.2018.12.015.

15. Zeng, T., and Li, Y.I. (2022). Predicting RNA splicing from DNA sequence using Pangolin. Genome Biol. 23, 103. 10.1186/s13059-022-02664-4.

16. Yeo, G., and Burge, C.B. (2004). Maximum Entropy Modeling of Short Sequence Motifs with Applications to RNA Splicing Signals. J. Comput. Biol. 11, 377–394. 10.1089/1066527041410418.

17. Shamsani, J., Kazakoff, S.H., Armean, I.M., McLaren, W., Parsons, M.T., Thompson, B.A., O’Mara, T.A., Hunt, S.E., Waddell, N., and Spurdle, A.B. (2019). A plugin for the Ensembl Variant Effect Predictor that uses MaxEntScan to predict variant spliceogenicity. Bioinformatics 35, 2315–2317. 10.1093/bioinformatics/bty960.

18. Eng, L., Coutinho, G., Nahas, S., Yeo, G., Tanouye, R., Babaei, M., Dörk, T., Burge, C., and Gatti, R.A. (2004). Nonclassical splicing mutations in the coding and noncoding regions of the ATM Gene: Maximum entropy estimates of splice junction strengths. Hum. Mutat. 23, 67–76. 10.1002/humu.10295.

19. Sudlow, C., Gallacher, J., Allen, N., Beral, V., Burton, P., Danesh, J., Downey, P., Elliott, P., Green, J., Landray, M., et al. (2015). UK Biobank: An Open Access Resource for Identifying the Causes of a Wide Range of Complex Diseases of Middle and Old Age. PLOS Med. 12, e1001779. 10.1371/journal.pmed.1001779.

20. Carss, K., Halldorsson, B.V., Hou, L., Liu, J., Wheeler, E., Lo, Y., Kundu, K., Huang, Z., Lacey, B., Dhindsa, R.S., et al. (2025). Whole-genome sequencing of 490,640 UK Biobank participants. Nature 645, 692–701. 10.1038/s41586-025-09272-9.

21. Sun, B.B., Chiou, J., Traylor, M., Benner, C., Hsu, Y.-H., Richardson, T.G., Surendran, P., Mahajan, A., Robins, C., Vasquez-Grinnell, S.G., et al. (2023). Plasma proteomic associations with genetics and health in the UK Biobank. Nature 622, 329–338. 10.1038/s41586-023-06592-6.

22. The 100,000 Genomes Project Pilot Investigators (2021). 100,000 Genomes Pilot on Rare-Disease Diagnosis in Health Care — Preliminary Report. N. Engl. J. Med. 385, 1868–1880. 10.1056/NEJMoa2035790.

23. Genomics England (2024). The National Genomic Research Library. 10.6084/m9.figshare.4530893.

24. Lord, J., Pagnamenta, A.T., Vestito, L., Walker, S., Oquendo, C.J., McGuigan, A.E., Ho, A., Odhams, C., Jacobsen, J.O., Mehta, S., et al. (2026). Blood-based RNA-Seq of 5412 individuals with rare disease identifies new candidate diagnoses in the National Genomic Research Library. Preprint at medRxiv, 10.64898/2026.03.19.26348811 https://doi.org/10.64898/2026.03.19.26348811.

25. Wright, C.F., Campbell, P., Eberhardt, R.Y., Aitken, S., Perrett, D., Brent, S., Danecek, P., Gardner, E.J., Chundru, V.K., Lindsay, S.J., et al. (2023). Genomic Diagnosis of Rare Pediatric Disease in the United Kingdom and Ireland. N. Engl. J. Med. 388, 1559–1571. 10.1056/NEJMoa2209046.

26. Foreman, J., Perrett, D., Mazaika, E., Hunt, S.E., Ware, J.S., and Firth, H.V. (2023). DECIPHER: Improving Genetic Diagnosis Through Dynamic Integration of Genomic and Clinical Data. Annu. Rev. Genomics Hum. Genet. 24, 151–176. 10.1146/annurev-genom-102822-100509.

27. McLaren, W., Gil, L., Hunt, S.E., Riat, H.S., Ritchie, G.R.S., Thormann, A., Flicek, P., and Cunningham, F. (2016). The Ensembl Variant Effect Predictor. Genome Biol. 17, 122. 10.1186/s13059-016-0974-4.

28. Karczewski, K.J., Francioli, L.C., Tiao, G., Cummings, B.B., Alföldi, J., Wang, Q., Collins, R.L., Laricchia, K.M., Ganna, A., Birnbaum, D.P., et al. (2020). The mutational constraint spectrum quantified from variation in 141,456 humans. Nature 581, 434–443. 10.1038/s41586-020-2308-7.

29. Morales, J., Pujar, S., Loveland, J.E., Astashyn, A., Bennett, R., Berry, A., Cox, E., Davidson, C., Ermolaeva, O., Farrell, C.M., et al. (2022). A joint NCBI and EMBL-EBI transcript set for clinical genomics and research. Nature 604, 310–315. 10.1038/s41586-022-04558-8.

30. Martin-Geary, A.C., Lecoquierre, F., Walker, S., Whiffin, N., and Dawes, R. (2025). Using SpliceAI to triage splice-altering variants in 7,220 individuals with rare conditions highlights limitations of the precomputed scores. Preprint at medRxiv, 10.1101/2025.08.27.25334471 https://doi.org/10.1101/2025.08.27.25334471.

31. Mudge, J.M., Carbonell-Sala, S., Diekhans, M., Martinez, J.G., Hunt, T., Jungreis, I., Loveland, J.E., Arnan, C., Barnes, I., Bennett, R., et al. (2025). GENCODE 2025: reference gene annotation for human and mouse. Nucleic Acids Res. 53, D966–D975. 10.1093/nar/gkae1078.

32. Goldfarb, T., Kodali, V.K., Pujar, S., Brover, V., Robbertse, B., Farrell, C.M., Oh, D.-H., Astashyn, A., Ermolaeva, O., Haddad, D., et al. (2025). NCBI RefSeq: reference sequence standards through 25 years of curation and annotation. Nucleic Acids Res. 53, D243–D257. 10.1093/nar/gkae1038.

33. Sullivan, P.J., Quinn, J.M.W., Wu, W., Pinese, M., and Cowley, M.J. (2024). SpliceVarDB: A comprehensive database of experimentally validated human splicing variants. Am. J. Hum. Genet. 111, 2164–2175. 10.1016/j.ajhg.2024.08.002.

34. Kircher, M., Witten, D.M., Jain, P., O’Roak, B.J., Cooper, G.M., and Shendure, J. (2014). A general framework for estimating the relative pathogenicity of human genetic variants. Nat. Genet. 46, 310–315. 10.1038/ng.2892.

35. Rentzsch, P., Witten, D., Cooper, G.M., Shendure, J., and Kircher, M. (2019). CADD: predicting the deleteriousness of variants throughout the human genome. Nucleic Acids Res. 47, D886–D894. 10.1093/nar/gky1016.

36. Hawkes, G., Chundru, K., Jackson, L., Patel, K.A., Murray, A., Wood, A.R., Wright, C.F., Weedon, M.N., Frayling, T.M., and Beaumont, R.N. (2025). Whole-genome sequencing analysis identifies rare, large-effect noncoding variants and regulatory regions associated with circulating protein levels. Nat. Genet. 57, 626–634. 10.1038/s41588-025-02095-4.

37. Benjamini, Y., and Hochberg, Y. (1995). Controlling the False Discovery Rate: A Practical and Powerful Approach to Multiple Testing. J. R. Stat. Soc. Ser. B Methodol. 57, 289–300. 10.1111/j.2517-6161.1995.tb02031.x.

38. Schymkowitz, J., Borg, J., Stricher, F., Nys, R., Rousseau, F., and Serrano, L. (2005). The FoldX web server: an online force field. Nucleic Acids Res. 33, W382–W388. 10.1093/nar/gki387.

39. Stephenson, J.D., Totoo, P., Burke, D.F., Jänes, J., Beltrao, P., and Martin, M.J. (2024). ProtVar: mapping and contextualizing human missense variation. Nucleic Acids Res. 52, W140–W147. 10.1093/nar/gkae413.

40. Tunyasuvunakool, K., Adler, J., Wu, Z., Green, T., Zielinski, M., Žídek, A., Bridgland, A., Cowie, A., Meyer, C., Laydon, A., et al. (2021). Highly accurate protein structure prediction for the human proteome. Nature 596, 590–596. 10.1038/s41586-021-03828-1.

41. Jaramillo Oquendo, C., Wai, H.A., Rich, W.I., Bunyan, D.J., Thomas, N.S., Hunt, D., Lord, J., Douglas, A.G.L., and Baralle, D. (2024). Identification of diagnostic candidates in Mendelian disorders using an RNA sequencing-centric approach. Genome Med. 16, 110. 10.1186/s13073-024-01381-w.

42. Al Masroori, E., Titheradge, H., Wai, H.A., Davies, I.C., Baralle, D., and Al-Abadi, E. (2026). A case report: camptodactyly-arthropathy-coxa vara-pericarditis syndrome: confirmation of a previously undescribed PRG4 splicing variant using synovial tissue RNA studies. EULAR Rheumatol. Open 2, 149–154. 10.1016/j.ero.2025.12.016.

43. Hoch, N.C., Hanzlikova, H., Rulten, S.L., Tétreault, M., Komulainen, E., Ju, L., Hornyak, P., Zeng, Z., Gittens, W., Rey, S.A., et al. (2017). XRCC1 mutation is associated with PARP1 hyperactivation and cerebellar ataxia. Nature 541, 87–91. 10.1038/nature20790.

44. O’Connor, E., Vandrovcova, J., Bugiardini, E., Chelban, V., Manole, A., Davagnanam, I., Wiethoff, S., Pittman, A., Lynch, D.S., Efthymiou, S., et al. (2018). Mutations in XRCC1 cause cerebellar ataxia and peripheral neuropathy. J. Neurol. Neurosurg. Psychiatry 89, 1230–1232. 10.1136/jnnp-2017-317581.

45. Abou Tayoun, A.N., Pesaran, T., DiStefano, M.T., Oza, A., Rehm, H.L., Biesecker, L.G., Harrison, S.M., and Svi), C.S.V.I.W.G. (ClinGen (2018). Recommendations for interpreting the loss of function PVS1 ACMG/AMP variant criterion. Hum. Mutat. 39, 1517–1524. 10.1002/humu.23626.

46. Fukuchi, S., Hosoda, K., Homma, K., Gojobori, T., and Nishikawa, K. (2011). Binary classification of protein molecules into intrinsically disordered and ordered segments. BMC Struct. Biol. 11, 29. 10.1186/1472-6807-11-29.

47. Gnanaolivu, R.D., and Hart, S.N. (2026). Enhancing missense variant classification in predicted intrinsically disordered regions. PLOS ONE 21, e0354365. 10.1371/journal.pone.0354365.

48. Wagner, N., Neverov, A.M., Martin-Geary, A.C., Londhe, S., Schröder, C., Yépez, V.A., Whiffin, N., and Gagneur, J. (2025). Aberrant splicing prediction during human organ development. Preprint at bioRxiv, 10.1101/2025.07.16.665183 https://doi.org/10.1101/2025.07.16.665183.

49. Cheng, J., Taylor, K.R., Nicolaisen, L., Pan, J., Bycroft, C., Perino, M., Ward, T., Hawkes, G., Covill, L.E., Weilert, M., et al. (2026). AlphaGenome Atlas: in silico mutagenesis of the entire human genome improves prioritization and interpretation of non-coding variants. Preprint at medRxiv, 10.64898/2026.09.16.26363192 https://doi.org/10.64898/2026.09.16.26363192.

