## Supplementary Figures for "Investigating the pathogenicity of missense spliceogenic variants using whole-genome sequencing, RNA-sequencing, and proteomics data from population and clinical research cohorts"

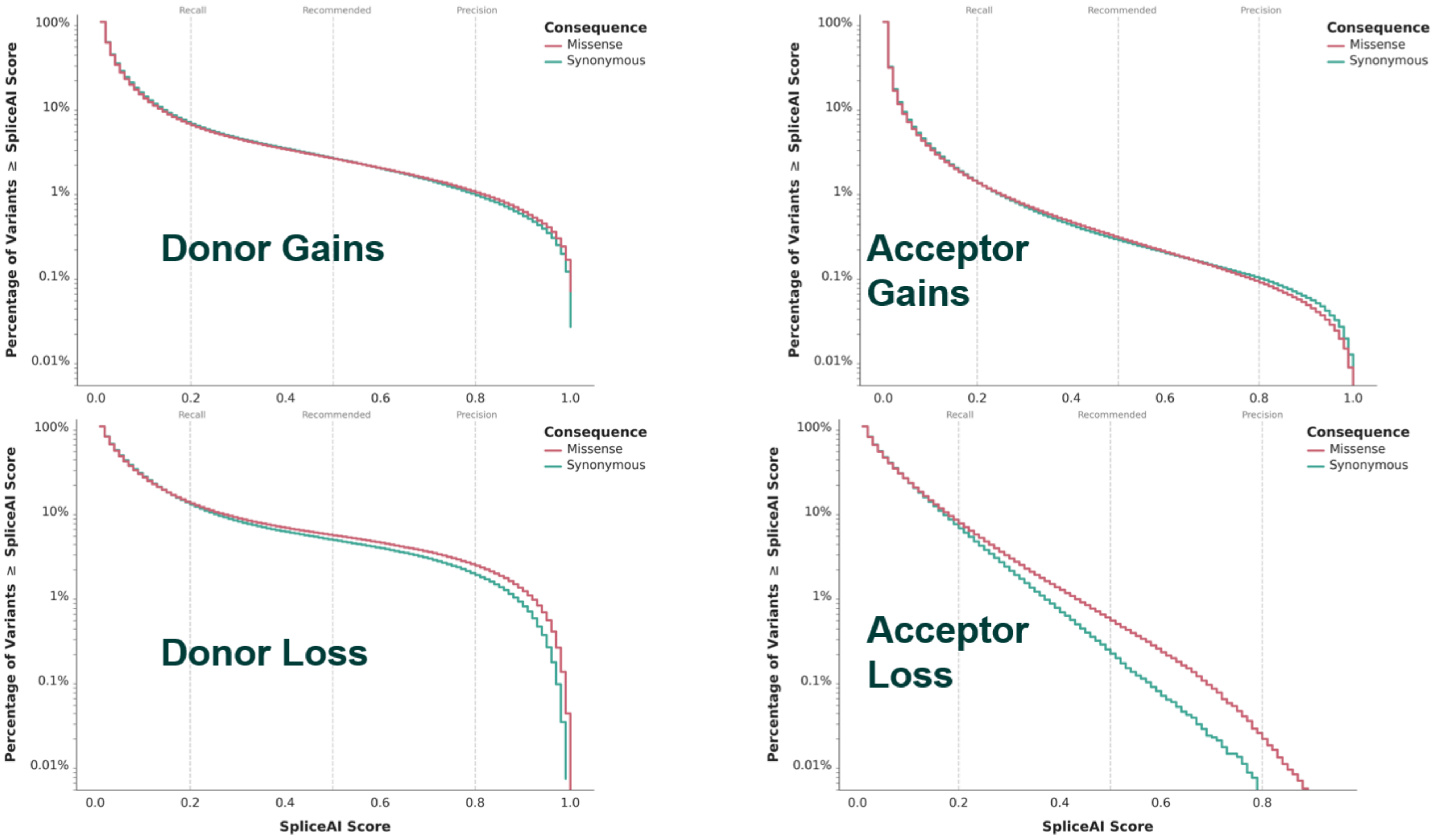


**Figure S1: Prevalence and spatial distribution of predicted missense spliceogenic variants (MSVs), stratified by splice event.**

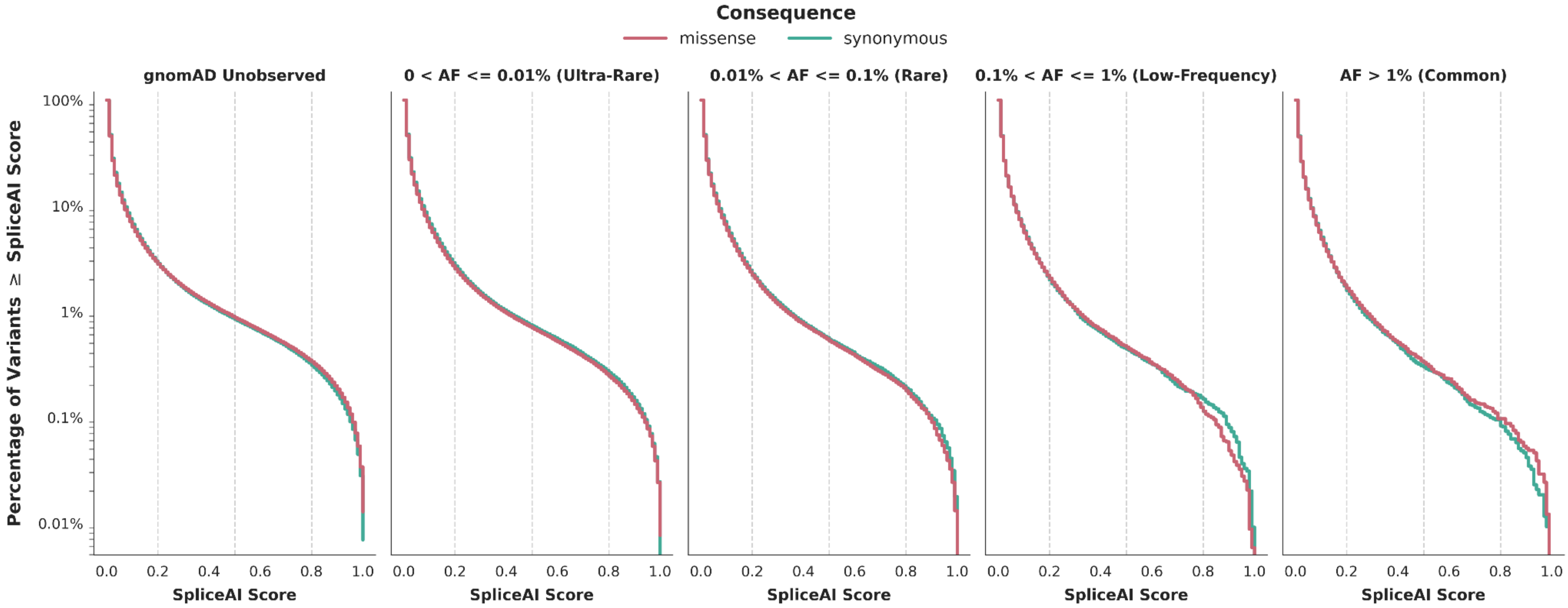


**Figure S2: Cumulative distribution of SpliceAIΔ scores for missense (red) and synonymous (teal) variants**. Vertical dashed lines denote standard thresholds for Recall (0.2), Recommended (0.5), and Precision (0.8) from SpliceAI. Stratified by gnomAD v4.1.1 Allele Frequency.

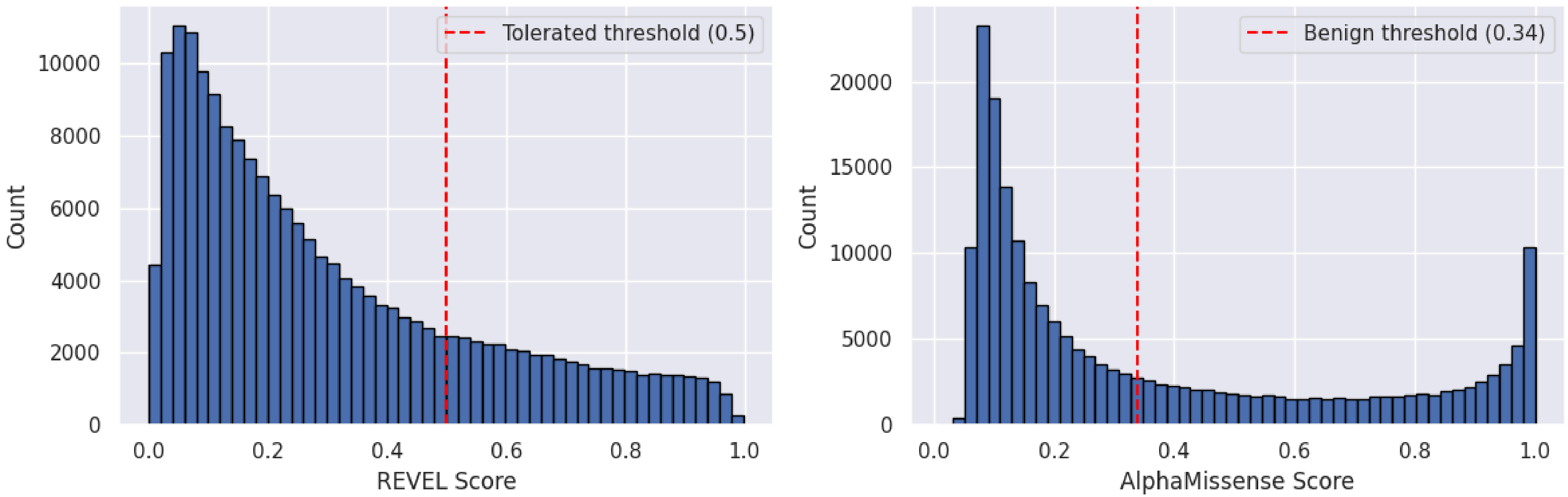


**Figure S3**: Distribution of REVEL and AlphaMissense scores for all UK Biobank missense variants with SpliceAI ≥ 0.2 (n = 225,430).

A)

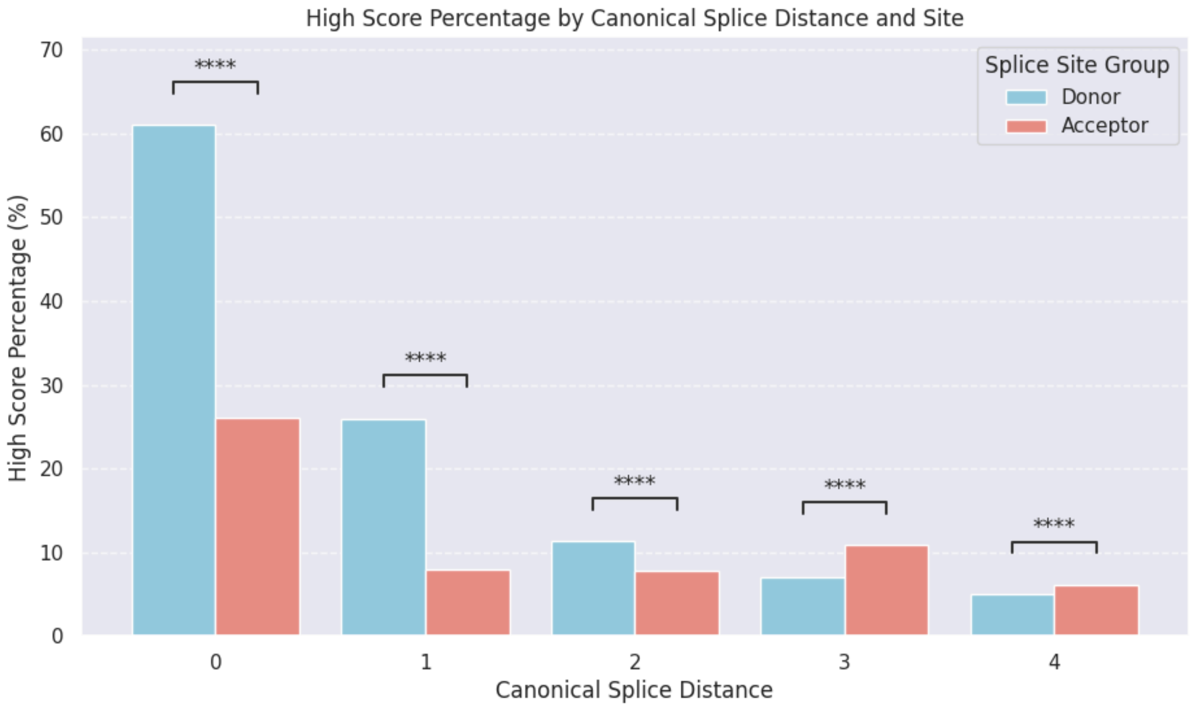


B)

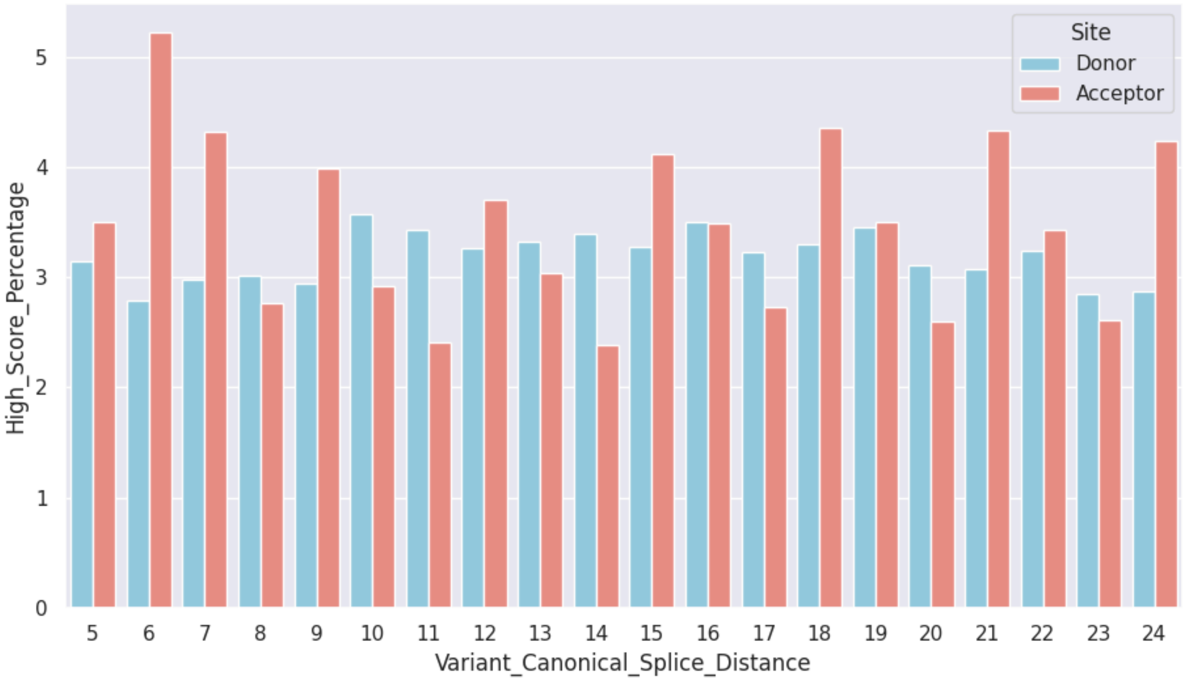


**Figure S4**: **Percentage of MANE Select UKB missense with a SpliceAIΔ of 0.2 or greater, binned by their distance to a canonical splice site and stratified by if in closer proximity to a donor or acceptor site.**

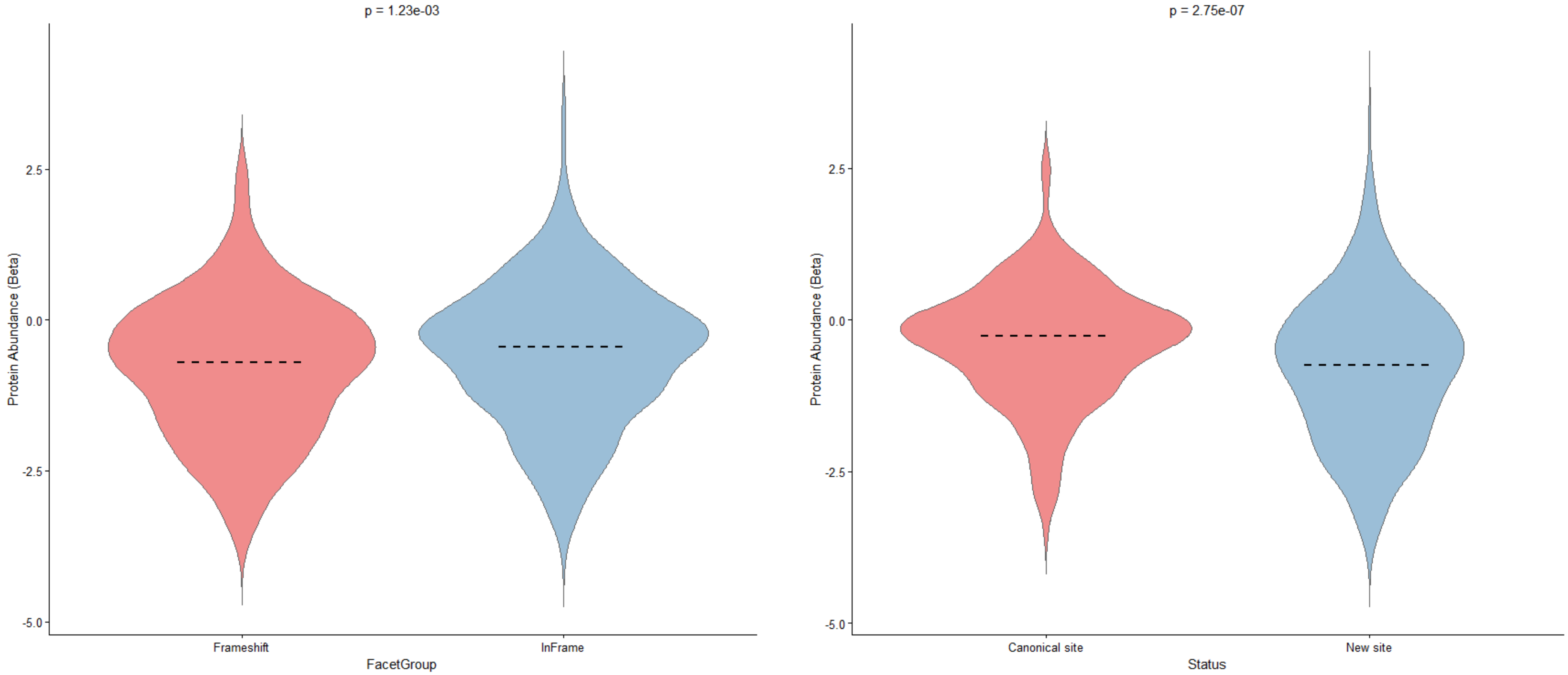


**Figure S5: Protein abundance grouped by SpliceAI predicted strongest junction and Frameshift event.**

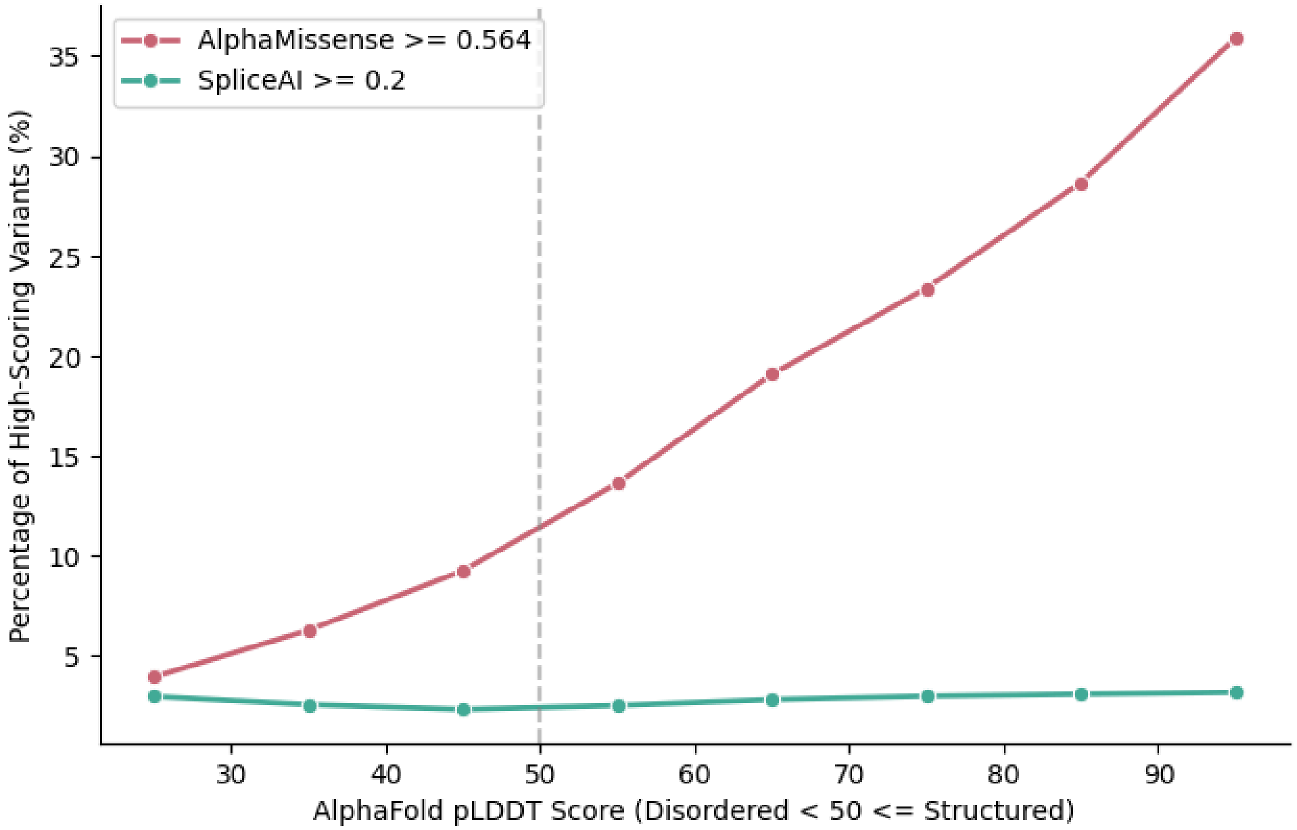


**Figure S6: Percentage of variants ≥ SpliceAIΔ 0.2 (green) and ≥ AlphaMissense 0.564 relative to the residue's pLDDT AlphaFold score.**
